# The impact of sexual violence in mid-adolescence on mental health: a UK population-based longitudinal study

**DOI:** 10.1101/2022.04.22.22274142

**Authors:** Francesca Bentivegna, Praveetha Patalay

## Abstract

**Background:** A large gender gap appears in internalising mental health during adolescence. There is little high-quality longitudinal population-based research investigating the role of sexual violence experiences, which are disproportionately experienced by females. This study aims to estimate the mental health impact of sexual violence experiences in mid-adolescence.

**Methods:** Longitudinal data from the UK Millennium Cohort Study (born 2000-02) in participants with information available on past-year sexual violence (sexual assault, unwelcome sexual approach) and mental health outcomes (psychological distress (K-6 questionnaire) in last 30 days, past-year self-harm, and lifetime attempted suicide) reported at age 17 years. Multivariable confounder adjusted regressions and propensity matching approaches were used, and population attributable fractions (PAFs) were calculated.

**Findings:** Analyses were in 5,119 girls and 4,852 boys (mean age 17 years, 80.8% White). In the fully adjusted model, sexual violence was associated with greater mean psychological distress (mean difference girls: 2.09 [1.51; 2.68] boys: 2.56 [1.59; 3.53]) and higher risk of high distress (girls: 1.65 [1.37; 2.00] boys: 1.55 [1.00; 2.40]), self-harming (girls: 1.79 [1.52; 2.10] boys: 2.16 [1.63; 2.84]), and attempted suicide (girls: 1.75 [1.26; 2.41] boys: 2.73 [1.59; 4.67]). PAF estimates suggest that, in a scenario with no sexual violence, we could expect 3.7-10.5% (boys) to 14.0-18.7% (girls) fewer adverse mental health outcomes at this age.

**Interpretation:** Our findings demonstrate the substantial role of sexual violence experiences for mid-adolescent mental health, especially for girls who are 4-5 times more likely to be victims. Changes are needed at societal and policy levels to prevent sexual violence and its wide-ranging impacts.

**Funding:** Medical Research Council.

## Introduction

There is a well-established and substantial gender gap in common mental illnesses, whereby women experience higher rates of depression, anxiety, and self-harm from adolescence onwards.^1,2^ There are many hypothesized explanations for this gender gap, including differential social risks, biological development, and specific vulnerability factors in interaction with stressors that are particularly salient in adolescence,^3,4^ and it is likely that several factors contribute to the larger burden of these mental health problems faced by women. One potential risk factor that is disproportionately experienced by females from early adolescence is sexual violence, including sexual abuse, assault, and harassment.^5^ Teenage girls experience over five times the levels of sexual assault compared to their male peers.^6^ Estimating the impact of sexual violence in mid-adolescence on subsequent mental health is, therefore, critical.^7^

The available evidence on the impact of sexual violence in mid-adolescence on mental health is scarce, as much attention pre-adulthood has been given to adverse childhood experiences, including sexual abuse earlier in childhood,^8,9^ experiences of sexual violence in the university and college environment,^10,11^ and intimate partner violence in older adolescent age groups.^12^ Several existing studies we identified that focused on mid-adolescence, an age at which many begin to experience increasing levels of sexual harassment and assault after puberty, are limited in their ability to make assumptions on the population level impacts of sexual violence due to either the use of selective samples, such as those recruited from sexual assault referral centres,^13^ the examination of cross-sectional associations,^14,15^ or the use of retrospective recollections of both violence experienced and its perceived subsequent impact on mental health.^7,16^ As for the studies with any longitudinal examination, these were limited to convenience samples in some North American school settings,^17,18^ and do not include relevant wider and earlier life factors that might be important confounders of this relationship.^19,20^

This paper aims to estimate the impact of sexual violence experienced in mid-adolescence on depressive symptoms, self-harm, and attempted suicide, in a prospective longitudinal cohort study. Although in many health contexts randomised control trials represent the gold standard for establishing causal impact, the ethical considerations for avoiding a design where sexual violence experiences are experimentally manipulated are clear. However, there are methods that allow the creation of a pseudo-experimental design, including propensity score matching (PSM).^21^ In this matching approach, each individual from the treatment group is matched to an individual in the control group based on similarity on pre-specified criteria. Due to the gendered nature of sexual violence experiences and different salience of these experiences in males and females,^18^ we present stratified analyses. We hypothesise that adolescents who experienced sexual violence will have worse subsequent mental health, and that effects will be robust to rich confounder adjustment and under the conditions of a pseudo-experimental design. Lastly, we estimate the population attributable fraction (PAF) to predict the reduction in adverse mental health outcomes if sexual violence at this age were successfully limited via necessary policy and societal changes.

## Methods

### Design

We used a longitudinal study with rich data on early life factors and possible confounders to estimate the impacts of sexual violence on mental health using two approaches: 1) multivariable regression with adjustment for a wide range of relevant confounders; 2) pseudo-experimental design with a group that experienced sexual violence, and a matched control group (Figure S1). Lastly, we combined these two methods by adjusting for a wider set of confounders in the context of the pseudo-experimental design.

### Participants

Data from a national birth cohort study, the Millennium Cohort Study, were used. The main analysis focused on data from two consecutive sweeps at age 14 (2015-16; 11,726 families) and age 17 (2018-19; 10,625 families),^22^ with confounders being from earlier sweeps at ages 3, 5, 7, 11 years as relevant.

For the multivariable analyses, participants had to have information available on an experience of sexual violence and one outcome. For the pseudo-experimental design with matched sexual violence-control group, the included participants were also required to have data on all the key matching variables (sample flowchart Figure 1).

**Figure 1.**
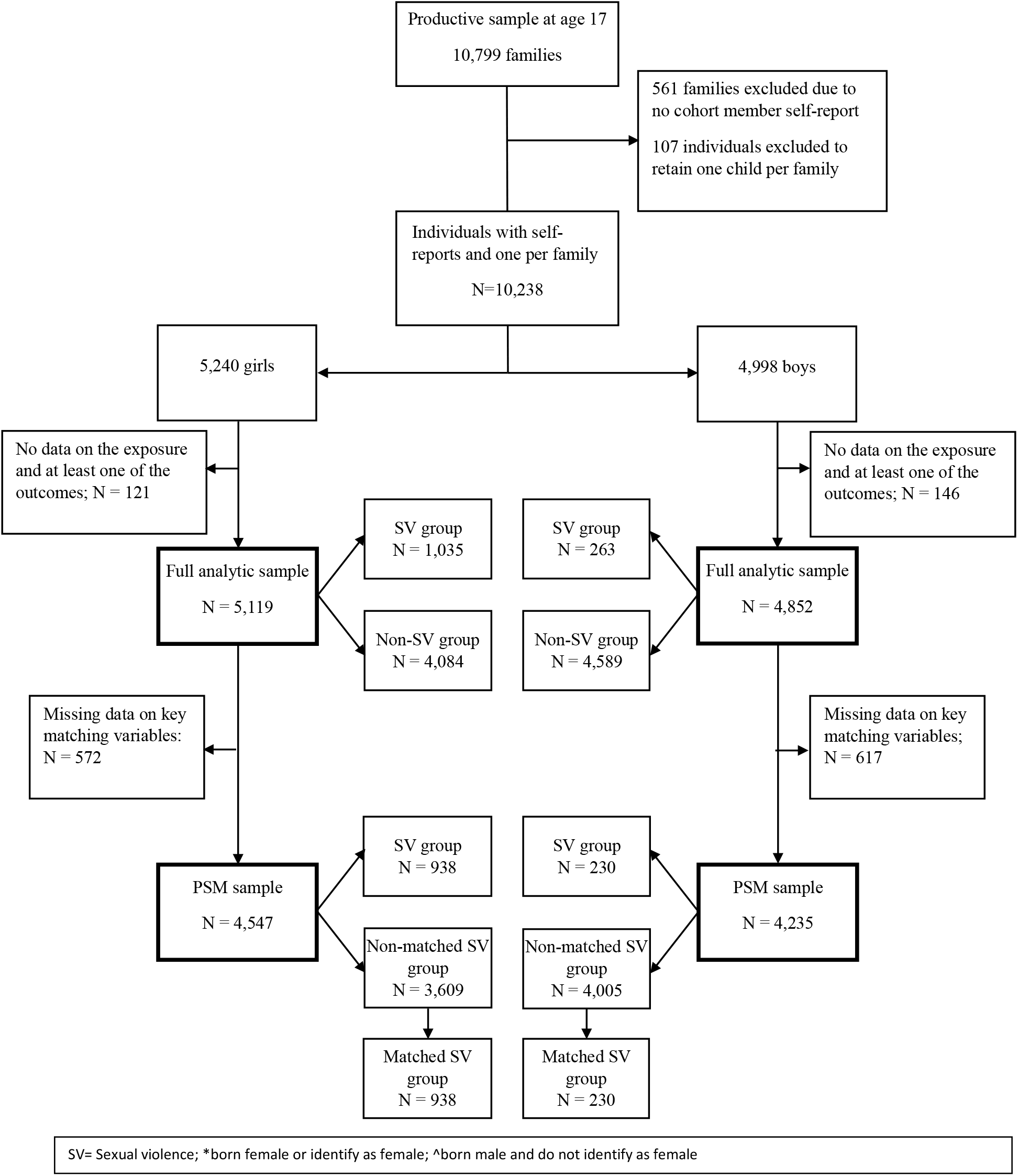
Sample flow diagram illustrating the analytic samples for the multivariable and PSM analyses

Ethical approvals for each sweep of MCS were received from National Research Ethics Committees (details in Supplement).

## Measures

### Sexual violence

*Sexual violence* was assessed using two questions from the self-reported questionnaire at age 17. Participants were asked about their experiences as victims of sexual assault and unwelcomed sexual approach in the previous 12 months with dichotomous (yes/no) responses. We created a binary sexual violence variable by combining information from these items, and also analysed them separately.

### Mental health outcomes

*Psychological distress* at age 17 was assessed using the Kessler-6 (K-6), a six-item, self-administered inventory measuring distress over the previous 30 days. Items refer to both depressive and anxiety symptoms and are rated on a 5-point Likert scale (‘all of the time’ to ‘none of the time’). A total score was obtained from summing the items, with scores of 13 or above indicating high distress. ^23^The measure was selected as it is the main measure capturing symptoms of common mental health conditions in the MCS cohort; included in the cohort because it is a well-validated short measure of population psychological distress with established clinical thresholds and used in many countries. Further details of the measure and its use are included in the Supplement.

*Self-harm* was assessed by asking participants whether (yes/no) they hurt themselves on purpose in the previous 12 months by cutting, burning, bruising, taking an overdose, pulling out hair, and hurting themselves in other ways. A binary variable was created indicating self-harming in the previous 12 months if participants responded affirmatively to any of these items.

*Attempted Suicide* was assessed via responses (yes/no) to the question ‘Have you ever hurt yourself on purpose in an attempt to end your life?’.

### Confounding variables

We chose the following matching and confounding factors in line with previous literature indicating the following as likely relevant confounders of sexual violence and mental health including prior mental health (depressive symptoms, self-harm), family and socio-demographic factors (ethnicity, sexuality, parent education, family income, number of siblings, number of parents/carers), puberty-related variables (age of menarche [only girls], pubertal status, early sexual activity, prior sexual violence); interpersonal relationships (relationship status, peer relationships, bullying); and health-related variables (BMI, risky behaviours, missing school without parents’ permission, disability, life satisfaction).

Detailed information regarding the justification and measurement of the confounding variables used are in the Supplement.

## Statistical analyses

All analyses were conducted using Stata v.17.

For missing information within the analytic sample, multiple imputation with chained equations was used with ten imputations for exposure, outcomes, and confounders, using *mi impute chained*. We used a combined sampling and non-response weight provided with MCS age 17 to account for sampling design and attrition from previous sweeps.

We conducted linear regressions for the continuous outcome (psychological distress) and Poisson regressions for the binary outcomes (high psychological distress, self-harm, and attempted suicide).

For each outcome we began with an unadjusted model, and this was followed by models with increasing levels of adjustment: first prior mental health problems (psychological distress and self-harm at age 14); then socio-demographic characteristics (ethnicity, parental education, family income, number of siblings, number of parents/carers, sexual orientation); puberty-related variables (age of menarche, pubertal status, early sexual activity, sexual violence at age 14); interpersonal relationships (relationship status, peer relationships, bullying); and lastly, in the fully adjusted model, we added health-related variables (BMI, risky behaviours, missing school without parents’ permission, disability, life satisfaction). The complex survey design of MCS was accounted for, and sample and attrition weights were used.

We used PSM^21^ to generate a control group that was matched to the sexual violence group on key variables (see Supplement for variables), using *psmatch2*^24^ and a randomly-ordered sample and optimal full matching to identify 1:1 closest match for each participant in the sexual violence group on the matching variables. The quality of the matching was assessed using covariate imbalance testing and presented using graphs. Once matched, simple unweighted linear and Poisson regressions were estimated to investigate the effect of group membership on outcomes. As a sensitivity analysis, matched control groups involving different matching variables were created to examine whether findings varied with differences in the matching variables used. We also conduct a sensitivity analysis with the PSM sample where we exclude those who report sexual violence before age 14 years to examine whether the estimates varied when focussing on a group who did not report prior sexual violence.

As a final robustness check, we combined both approaches and merged the matched PSM groups with the full imputed sample in order to adjust the analysis in the matched sample for all the additional confounders not accounted for by matching.

As an exploratory analysis, we conducted multivariate analyses separately for unwelcome sexual approach and sexual assault to evaluate whether the observed effect sizes were similar for these exposures considered separately.

Lastly, we estimated PAFs to provide an interpretable estimate of effect sizes^25^ using punaf^26^ for the overall sexual violence exposure, and for unwelcome sexual approach and sexual assault, separately. The PAF defines the fraction of all cases that is attributable to a specific exposure and can be used to estimate the percentage of a certain outcome in a hypothetical scenario where the exposure is null.

The funder had no role in study design, data collection, data analysis, data interpretation, or writing of the report.

## Results

This study was conducted between 1^st^ February 2021 and 21^st^ June 2022.

Figure 1 shows the flow diagram and sample selection for the different analyses in girls (full sample n = 5,119, White = 4,138, Black = 181, Asian = 561, Mixed = 159, Other = 80) and boys (full sample n = 4,852, White = 3,925, Black = 178, Asian = 528, Mixed = 136, Other = 84).

Table 1 shows the weighted numbers and percentages, and means and standard deviations, for the full analytic sample (sexual violence group vs non-sexual violence group), and Table S1 shows these for the PSM sample (sexual violence group vs matched control group). Figure 2 displays the descriptive prevalence for the matched groups at ages 14 and 17 years.

**Table 1.** Descriptive analysis of mental health outcomes in different analytic samples.

|  |  | Girls |  |  | Boys |  |  |
| --- | --- | --- | --- | --- | --- | --- | --- |
|  |  | Full analytic sample<br>(N = 5,119) | Full analytic sample:<br>Sexual violence<br>group (N = 1,035) | Full analytic sample:<br>Non-sexual violence<br>group (N = 4,084) | Full analytic sample<br>(N = 4,852) | Full analytic sample:<br>Sexual violence<br>group (N = 263) | Full analytic sample:<br>Non-sexual violence<br>group (N = 4,589) |
|  |  | N (%) or M (SE)<br>[95% CI] | N (%) or M (SE)<br>[95% CI] | N (%) or M (SE)<br>[95% CI] | N (%) or M (SE) [95%<br>CI] | N (%) or M (SE) [95%<br>CI] | N (%) or M (SE) [95%<br>CI] |
| **Mental health at age 17**<br>**(reference period)** |  |  |  |  |  |  |  |
| Psychological distress<br>(30 days) | (continuous) | 8.41 (0.18)<br>[8.06, 8.75] | 10.92 (0.25) –<br>[10.43, 11.42] | 7.75 (0.21) –<br>[7.33, 8.17] | 6.17 (0.12) –<br>[5.95, 6.40] | 9.72 (0.34) –<br>[9.04, 10.39] | 5.97 (0.12) –<br>[5.73, 6.20] |
| High psychological<br>distress (binary)<br>(30 days) | No | 3,974 (77.4)<br>[74.9, 79.7] | 644 (60.0) –<br>[55.0, 65.3] | 3,330 (81.8) –<br>[79.0, 84.3] | 4,376 (89.8) –<br>[88.0, 91.4] | 192 (77.2) –<br>[67.5, 84.6] | 4,184 (90.5) –<br>[88.7, 92.1] |
|  | Yes | 1,141 (22.6)<br>[20.3, 25.1] | 390 (40.0) –<br>[34.7, 45.0] | 751 (18.2) –<br>[15.7, 21.0] | 474 (10.2) –<br>[8.6, 12.0] | 71 (22.8) –<br>[15.4, 32.5] | 403 (9.5) –<br>[7.9, 11.3] |
| Self-harm<br>(12 months) | No | 3,520 (71.7)<br>[69.3, 73.9] | 475 (47.3) –<br>[42.8, 51.9] | 3,045 (77.6) –<br>[75.1, 80.0] | 3,932 (79.9) –<br>[77.3, 82.3] | 134 (44.3) –<br>[31.4, 58.1] | 3,798 (81.9) –<br>[79.5, 84.1] |
|  | Yes | 1,436 (28.3)<br>[26.1, 30.7] | 506 (52.7) –<br>[48.1, 57.2] | 930 (22.4) –<br>[20.0, 24.9] | 826 (20.1) –<br>[17.7, 22.7] | 116 (55.7) –<br>[41.9, 68.6] | 710 (18.1) –<br>[15.9, 20.5] |
| Attempted suicide<br>(lifetime) | No | 4,437 (89.4)<br>[87.8, 90.71] | 791 (78.2) –<br>[73.0, 82.5] | 3,646 (92.1) –<br>[90.7, 93.3] | 4,551 (95.8) –<br>[94.8, 96.6] | 207 (85.1) –<br>[77.0, 90.7] | 4,344 (96.4) –<br>[95.4, 97.1] |
|  | Yes | 509 (10.6)<br>[9.3, 12.2] | 188 (21.8) –<br>[17.46, 27.0] | 321 (7.9) –<br>[6.7, 9.3] | 209 (4.2) –<br>[3.4, 5.2] | 44 (14.9) –<br>[9.3, 23.0] | 165 (3.6) –<br>[2.9, 4.6] |
| **Mental health at age 14**<br>**(reference period)** |  |  |  |  |  |  |  |
| Depressive symptoms<br>(2 weeks) | (continuous) | 7.10 (0.14) –<br>[6.82, 7.38] | 9.60 (0.36) –<br>[8.88, 10.31] | 6.44 (0.14) –<br>[6.16, 6.72] | 4.05 (0.11) –<br>[3.84, 4.26] | 5.55 (0.47) –<br>[4.62, 6.47] | 3.97 (0.11) –<br>[3.75, 4.19] |
| High depressive<br>symptoms (SMFQ ≥ 12)<br>(2 weeks) | No | 3,599 (77.5)<br>[75.9, 79.1] | 609 (64.1)<br>[59.9, 68.1] | 2,990 (81.1)<br>[79.4 82.7] | 3,972 (91.2) –<br>[89.8, 92.4] | 196 (86.0) –<br>[80.3, 90.2] | 3,776 (91.5) –<br>[90.0, 92.8] |
|  | Yes | 1,038 (22.5)<br>[20.9, 24.1] | 344 (35.9)<br>[31.8, 40.1] | 694 (18.9)<br>[17.3, 20.6] | 352 (8.8) –<br>[7.6, 10.2] | 41 (14.0) –<br>[9.8, 19.7] | 311 (8.5) –<br>[7.2, 10.0] |
| Self-harm<br>(12 months) | No | 3,648 (77.9) –<br>[76.1, 79.6] | 605 (63.8) –<br>[59.2, 68.1] | 3,043 (81.6) –<br>[79.8, 83.3] | 3,991 (91.0) –<br>[89.2, 92.4] | 190 (82.6) –<br>[76.8, 87.2] | 3,801 (91.4) –<br>[89.6, 92.9] |
|  | Yes | 993 (22.1) –<br>[20.4, 23.9] | 344 (36.2) –<br>[31.9, 40.8] | 649 (18.4) –<br>[16.7, 20.2] | 356 (9.0) –<br>[7.6, 10.8] | 49 (17.4) –<br>[12.8, 23.2] | 307 (8.6) –<br>[7.1, 10.4] |

N numbers; % percentage; M mean; SD standard deviation; CI confidence interval. Note: These are weighted using a combined sample and non-response weight for the analytic sample.

**Figure 2.**
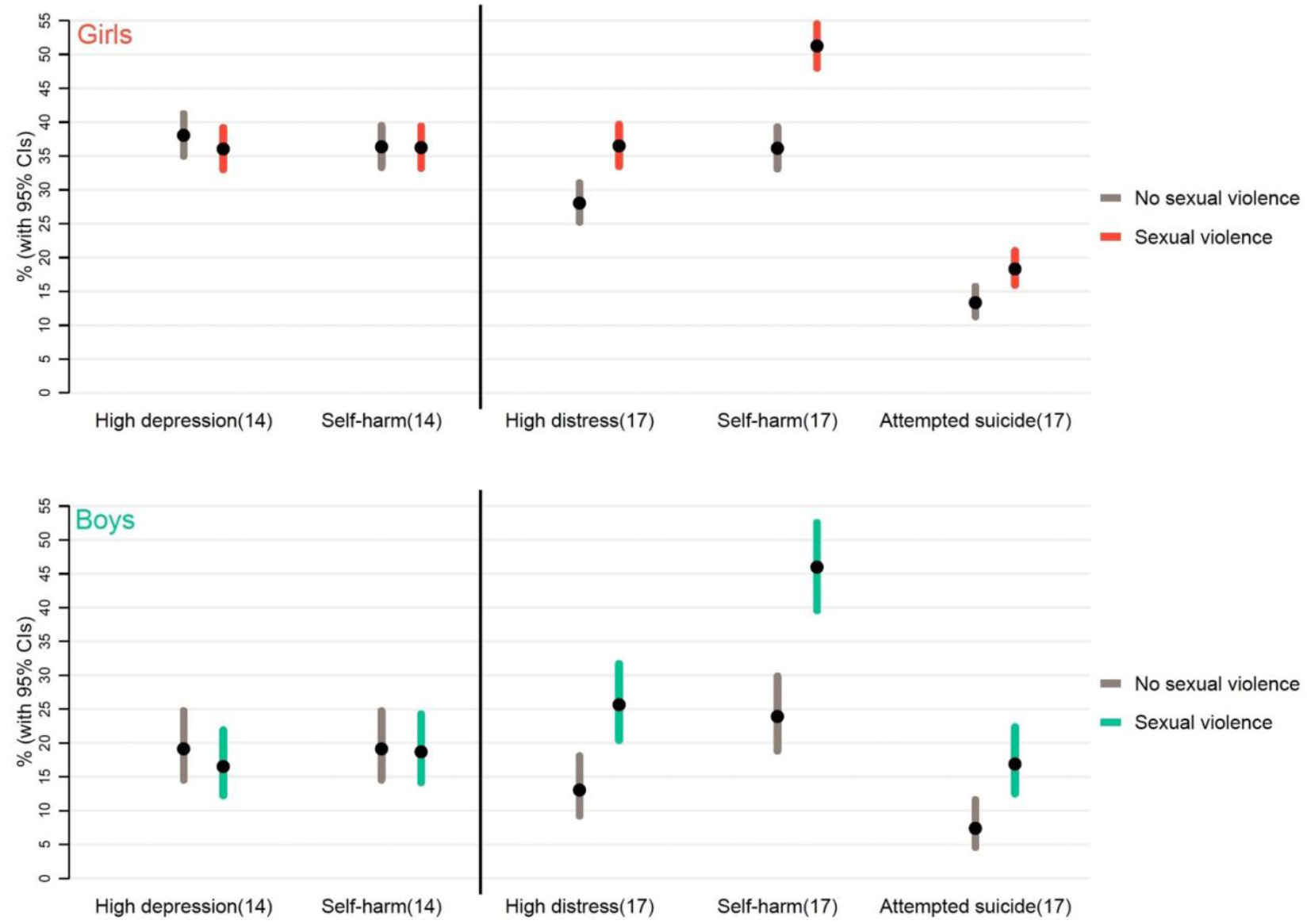
Prevalence of high depressive symptoms, self-harm, and attempted suicide at age 14 and at age 17 in the group that experienced sexual violence between these ages compared to the group that did not experience sexual violence (top panel girls, bottom panel boys). Note. This graph is based on prevalence in propensity score matched groups. N in each sexual violence group: girls = 938, boys = 225. Prevalence of high depression/distress are not comparable across ages 14 and 17. Depressive symptoms at 14 assesses using the Short Moods and Feelings Questionnaire and distress at age 17 using the Kessler K-6 measure of psychological distress.

**Figure 3.**
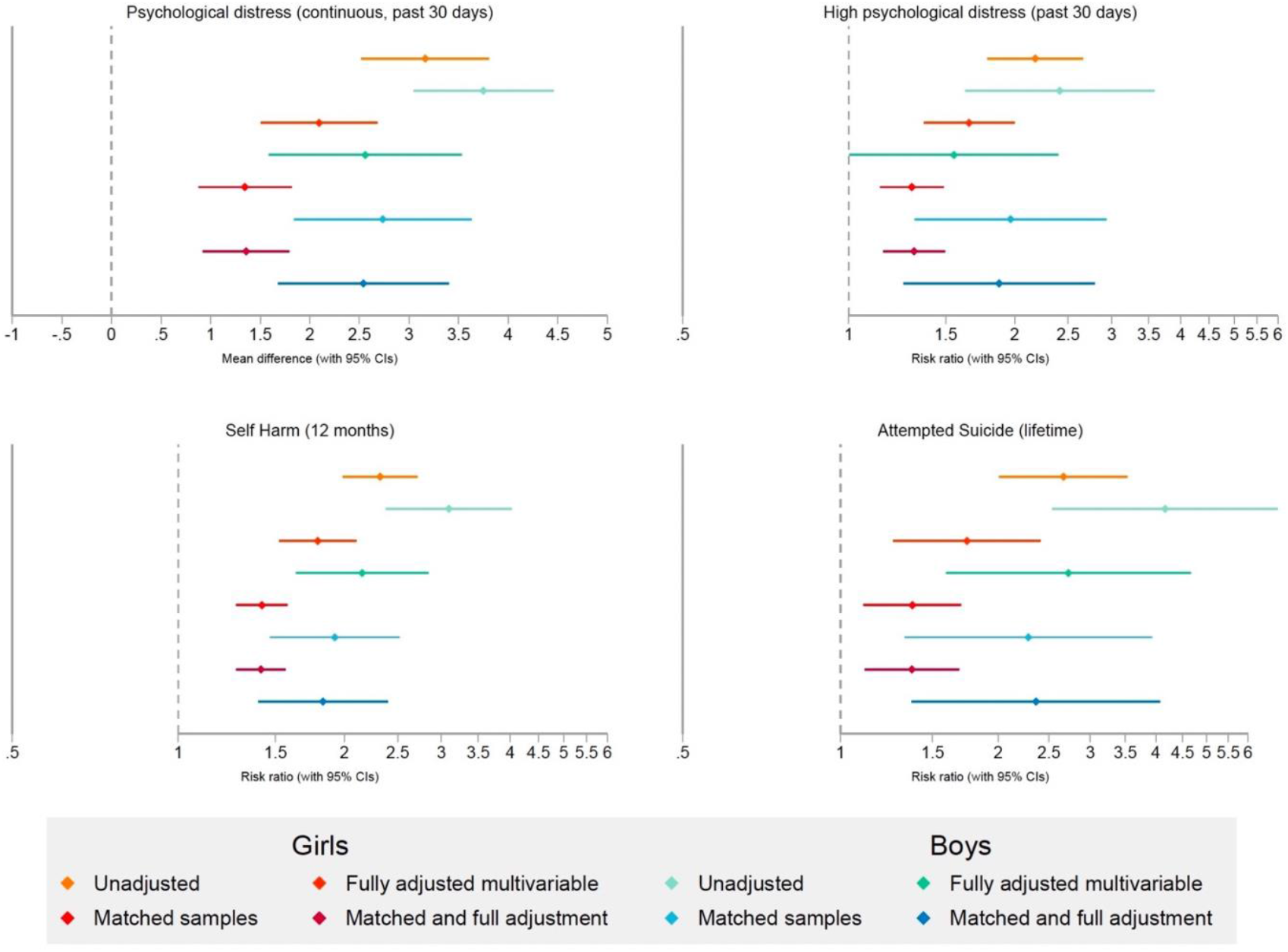
Multivariable regression analyses and PSM analyses estimating the impact of sexual violence on psychological distress (as both a continuous and binary measure), self-harm, and attempted suicide at age 17.

In girls, 20.2% (1,035/5,119) experienced any sexual violence (5.3% assault [269/5,119]; 19.4% unwelcome sexual approach [991/5,119]). In boys, 5.4% [263/4,852] experienced any sexual violence (1.0% [50/4,852] sexual assault; 5.2% [251/4,852] unwelcome sexual approach). Venn diagrams (Figure S2) show overlap between the two sub-types and highlight that most participants who reported assault also reported unwelcome sexual approaches.

Multivariable regressions in the multiply imputed full analytic sample with increasing levels of adjustment indicate that experiencing sexual violence prior to age 17 years was associated with worse mental health-outcomes at age 17 (Table 2). Specifically, in the final adjusted model, having experienced sexual violence was associated with a higher mean psychological distress (mean difference girls: 2.09 [1.51; 2.68], boys: 2.56 [1.59; 3.53]), and with a higher risk of experiencing high psychological distress (RR girls: 1.65 [1.37; 2.00], boys: 1.55 [1.00; 2.40]), self-harming (girls: 1.79 [1.52; 2.10]; boys: 2.16 [1.63; 2.84]), and attempting suicide (girls: 1.75 [1.26; 2.41]; boys: 2.73 [1.59; 4.67]). Complete case analyses are presented in Table S2 and show broadly similar results with bias in the expected direction (i.e., underestimation) due to attrition being predicted by socio-economic and other disadvantages, and poorer health.

**Table 2.** Regression coefficients and risk ratios for sexual violence predicting mental health outcomes from a range of modelling approaches.

|  | Psychological distress (continuous)<br>(30 days)<br>Coefficient [95% CI] |  | High psychological distress<br>(30 days)<br>RR [95% CI] |  | Self-harm<br>(12 months)<br>RR [95% CI] |  | Attempted Suicide<br>(lifetime)<br>RR [95% CI] |  |
| --- | --- | --- | --- | --- | --- | --- | --- | --- |
|  | Girls | Boys | Girls | Boys | Girls | Boys | Girls | Boys |
| **Multivariable regressions** |  |  |  |  |  |  |  |  |
| Unadjusted | 3.16 [2.52, 3.81] | 3.75 [3.04, 4.46] | 2.18 [1.79, 2.66] | 2.41 [1.62, 3.59] | 2.32 [1.99, 2.71] | 3.09 [2.38, 4.03] | 2.67 [2.01, 3.54] | 4.17 [2.54, 6.84] |
| Adjustment 1 | 2.14 [1.47, 2.81] | 3.11 [2.18, 4.05] | 1.67 [1.36, 2.05] | 1.87 [1.23, 2.84] | 1.90 [1.60, 2.25] | 2.62 [1.90, 3.62] | 1.84 [1.32, 2.56] | 3.00 [1.84, 4.88] |
| Adjustment 2 | 2.02 [1.37, 2.67] | 2.59 [1.61, 3.56] | 1.62 [1.32, 1.98] | 1.58 [1.02, 2.45] | 1.79 [1.52, 2.09] | 2.23 [1.74, 2.87] | 1.72 [1.22, 2.41] | 2.80 [1.66, 4.72] |
| Adjustment 3 | 2.10 [1.45, 2.75] | 2.57 [1.60, 3.54] | 1.65 [1.35, 2.02] | 1.57 [1.01, 2.44] | 1.81 [1.54, 2.12] | 2.22 [1.72, 2.88] | 1.74 [1.24, 2.43] | 2.60 [1.53, 4.41] |
| Adjustment 4 | 2.07 [1.44, 2.71] | 2.64 [1.69, 3.59] | 1.64 [1.35, 1.99] | 1.64 [1.08, 2.50] | 1.80 [1.54, 2.11] | 2.24 [1.73, 2.91] | 1.75 [1.26, 2.44] | 2.60 [1.55, 4.36] |
| Fully adjusted model | 2.09 [1.51, 2.68] | 2.56 [1.59, 3.53] | 1.65 [1.37, 2.00] | 1.55 [1.00, 2.40] | 1.79 [1.52, 2.10] | 2.16 [1.63, 2.84] | 1.75 [1.26, 2.41] | 2.73 [1.59, 4.67] |
| **Matched samples** |  |  |  |  |  |  |  |  |
| PSM (all matching variables) | 1.35 [0.88, 1.81] | 2.73 [1.84, 3.63] | 1.30 [1.14, 1.49] | 1.97 [1.31, 2.93] | 1.42 [1.28, 1.58] | 1.92 [1.47, 2.52] | 1.37 [1.11, 1.70] | 2.28 [1.33, 3.93] |
| PSM (all minus sexual identity, pubertal status, parental education, and sexual violence <14) | 1.66 [1.19, 2.14] | 3.29 [2.41, 4.16] | 1.35 [1.18, 1.54] | 2.68 [1.70, 4.23] | 1.58 [1.41, 1.77] | 2.18 [1.64, 2.92] | 1.44 [1.16, 1.79] | 4.28 [2.12, 8.65] |
| PSM (all minus psychological distress, self-harm at age 14, and sexual violence <14) | 2.37 [1.92, 2.83] | 3.53 [2.67, 4.38] | 1.72 [1.48, 2.00] | 3.11 [1.91, 5.04] | 1.82 [1.62, 2.06] | 2.24 [1.67, 3.00] | 2.17 [1.69, 2.80] | 4.83 [2.30, 10.14] |
| **Matched sample with multivariable confounder adjustment** |  |  |  |  |  |  |  |  |
| Adjusted regression analysis | 1.35 [0.92, 1.79] | 2.54 [1.68, 3.40] | 1.31 [1.16, 1.49] | 1.87 [1.26, 2.80] | 1.41 [1.27, 1.57] | 1.83 [1.40, 2.40] | 1.37 [1.11, 1.68] | 2.36 [1.37, 4.08] |
| **Multivariable adjusted regression by sexual violence type** |  |  |  |  |  |  |  |  |
| Unwelcomed sexual approach | 2.03 [1.44, 2.63] | 2.61 [1.61, 3.60] | 1.49 [1.34, 1.65] | 1.88 [1.44, 2.45] | 1.56 [1.44, 1.70] | 1.99 [1.67, 2.37] | 1.44 [1.21, 1.71] | 2.54 [1.72, 3.76] |
| Sexual assault | 2.32 [1.49, 3.15] | 3.82 [1.82, 5.81] | 1.46 [1.27, 1.68] | 1.87 [1.11, 3.13] | 1.57 [1.41, 1.75] | 2.09 [1.56, 2.80] | 1.72 [1.40, 2.11] | 2.63 [1.38, 5.01] |
| Unwelcome sexual approach only* | 1.51 [1.16, 1.86] | 2.05 [1.49, 2.60] | 1.45 [1.29, 1.64] | 1.80 [1.36, 2.38] | 1.49 [1.35, 1.65] | 1.87 [1.54, 2.26] | 1.27 [1.03, 1.57] | 2.33 [1.55, 3.53] |

RR risk ratio; CI confidence interval. All $p < 0.0001$ . PSM= Propensity score matched. \* Those who reported unwelcome sexual approach and no sexual assault
Adjustment 1: prior mental health (depressive symptoms and self-harm until 14 years). Adjustment 2: sociodemographic/economic characteristics (ethnicity, parental education, family income, sexual identity, single parent household, number of siblings). Adjustment 3: puberty-related characteristics (pubertal status, age of menarche (girls), early sexual activity, sexual violence until 14 years). Adjustment 4: interpersonal characteristics (relationship status, peer relationships, bullying). Fully adjusted model: health-related characteristics (BMI, risky behaviours, missing school without parents' permission, disability, life satisfaction). As can be seen in the table there is a substantial impact on the estimates of some confounder adjustment with later adjustment having less impact on the estimates.

As shown in Table S3a, S3b, and S4, the percentages and means for exposure, outcomes, and each of the key matching variables, were very similar in the full sample and in the PSM sample (for a comparison of the matching variables before and after matching see Table S5). The quality of the matching was assessed through evaluation of the percentage of bias for the exposure, outcomes, and matching variables (see Tables S6a and S6b; Figures S3a and S3b). Such bias was found to be low (<5%) for most variables, with a larger bias up to 10.3% for variables including other ethnic group, depressive symptoms at 14 years, and sexual violence prior to age 14.

In the PSM matched samples, the sexual violence group, on average, scored higher on psychological distress compared to the matched control group (Table 2; mean difference girls: 1.35 [0.88; 1.81], boys: 2.73 [1.84; 3.63]), and had greater risk of high distress (girls: 1.30 [1.14; 1.49], boys = 1.97 [1.31; 2.93]), self-harm (girls: 1.42 [1.28; 1.58] boys: 1.92 [1.47; 2.52]), and attempted suicide (girls: 1.37 [1.11; 1.70], boys: 2.28 [1.33; 3.93]).

As sensitivity analyses for the PSM, we used different matching variables to evaluate the best match as well as the difference in their estimates. Slightly different effect sizes were estimated based on the variables included in the matching, with often higher estimates when the matching was conducted with fewer variables (Table 2). Sensitivity analyses matching a sample excluding those who had reported experiences of sexual violence before age 14 demonstrate similar effect sizes to the analyses in the full sample (Table S7).

Analyses combining the matching and the full confounder adjustment resulted in similar effect sizes (Table 2), suggesting that the estimates are likely to be robust and any unmeasured confounder would have to be fairly strong to explain away observed effects.

Additionally, we run multivariate analyses for unwelcomed sexual approach, sexual assault, and unwelcome sexual approach only (excluding the subset who also experienced sexual assault), as seen in Table 2.

Finally, we estimated PAFs, which are based on the assumption that the estimates are causal (Table S8; Figure 4). In a scenario where girls did not experience sexual violence, there would be approximately 14.0-18.7% fewer serious mental health problems – e.g., translating to 9.1% prevalence of attempted suicide (compared to 11.0% observed). For boys, there would be approximately 3.7-10.5% fewer mental health problems – e.g., translating to 3·8% prevalence of attempted suicide (compared to 4.3% observed).

**Figure 4.**
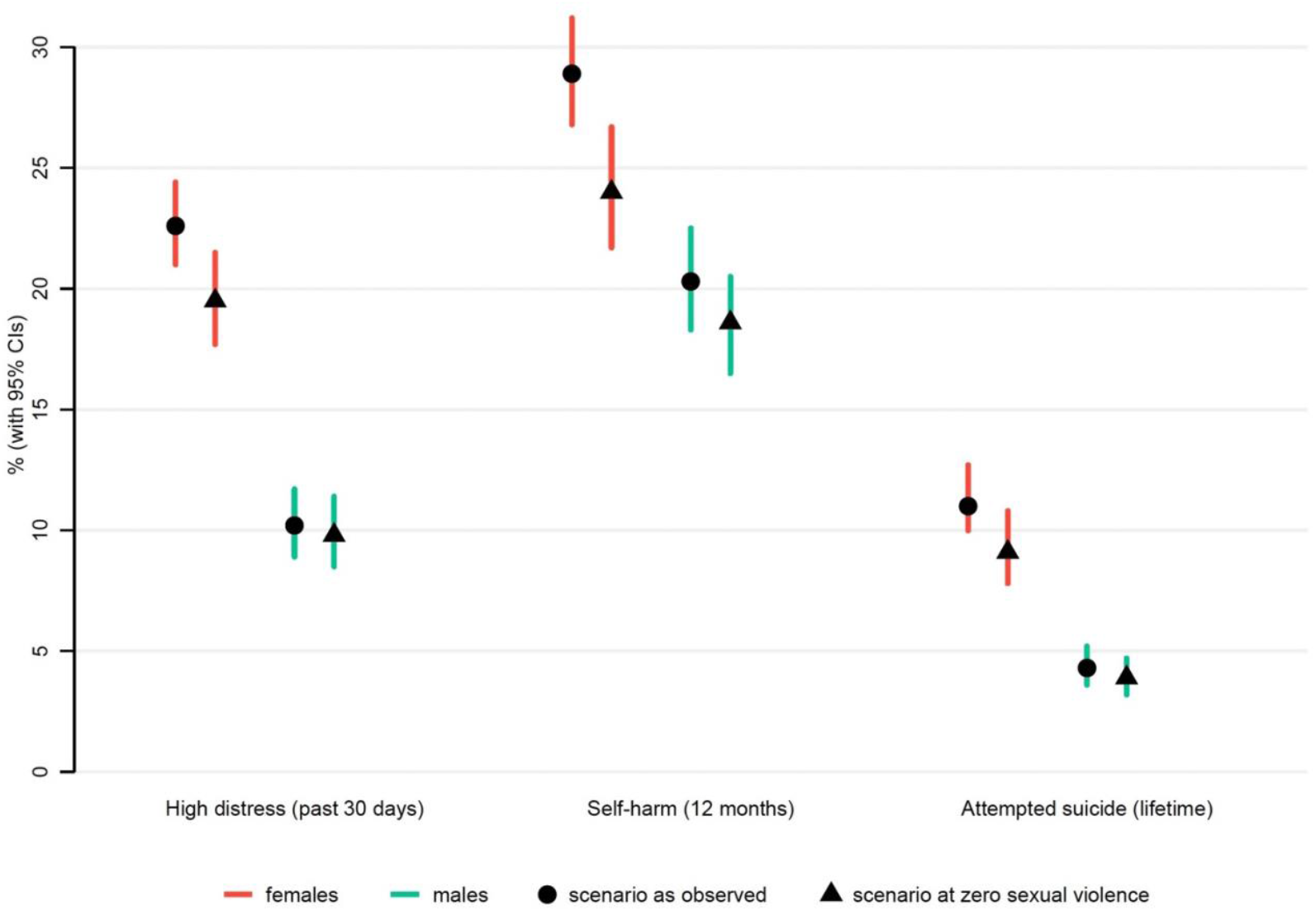
Results from the Population Attributable Fraction analysis estimating observed prevalence of the outcomes at age 17 years, and prevalence in a hypothetical counterfactual scenario where there were no sexual violence experiences at this age.

## Discussion

In our sample, adolescents who were victims of sexual violence in the year before age 17 years reported worse mental health outcomes at age 17 years, compared to those without these experiences. These effects persisted even after accounting for previous psychological distress and self-harm, and a wide range of relevant confounders, and were robust to multiple methodological approaches and sensitivity checks.

Because this study was based on data from the general population, the reported effect sizes can be considered in a public health context as they apply to the population. Due to the sizes of effect being large for all the analysed mental health outcomes (RRs > 1.30), these findings hold potentially important real-world implications. Our PAF findings, assuming this effect is causal and robust to further potential confounding, suggest that the absence of these experiences at this age would result in a 14.0-18.7% relative reduction in girls’ and 3.7-10.5% in boys’ mental health adverse outcome prevalence (high distress prevalence in girls 19.5% instead of 22.6% observed; in boys 9.8% instead of 10.2% observed), thus also substantially narrowing the gender gap reported at this age.^2^

In other words, sexual violence at this age contributes non-trivially to the large prevalence of mental ill-health observed, especially in adolescent girls, and actively tackling sexual violence could entail significant improvements in adolescent mental health. In a recent report in the UK, estimates from school populations showed high levels of sexual violence, with 79% of the surveyed girls reporting sexual assault of any kind.^27^ The same report highlighted the perceived inadequacy of school-based sex education. However, peer-on-peer in schools could be tackled with appropriate school-based intervention and education.^28^ This is even more relevant when considering the high prevalence of adolescents, especially girls, with a history of sexual violence and the fact that many do not report such experiences due to the stigma around these problems.^29^ Other avenues for intervention include within families and interpersonal relationships via social services, primary care, and other community-based services.^30^

Many factors, from biological, social to familial, and ascertainment biases, have been attributed as causes for the emergent gender gap in depression and other mental health difficulties in adolescence.^1,3^ For instance, greater pressures around body image and academic achievements are often cited as important gendered risk factors. Our findings clearly highlight the role of sexual violence, that is disproportionally experienced by girls, as an important and preventable driver of the gender gap in adolescent mental health.

The strengths of this study included a population-based cohort, longitudinal data, and adjustment for a rich set of important factors that might confound this association. These strengths of the data, accompanied with methods that aim to improve causal inference, help provide robust estimates of the mental health impacts of sexual violence in adolescence. However, there were also some limitations. First, information on mental health and prior sexual violence were collected at the same time in the age 17 assessment, and there is some evidence that mental health can be a predictor in the recall and reporting of these adverse experiences.^31^ The self-harm with suicidal intention question at age 17 asked about lifetime prevalence (ever), and although we control for self-harming before age 14 years, there are issues around timing and temporal precedence of exposure and outcome that we cannot resolve. In addition, reported prevalence of violence experiences, albeit high, is lower than suggested by some other data sources.^5,27^ Hence, it is possible that the association we observe is overestimated as adolescents with fewer mental health difficulties might not have reported their sexual violence experiences; on the other hand, it is possible that underreporting might have resulted in downward bias and an underestimation of effects. Future studies with more and better longitudinal follow-up data will allow further unpacking of these findings, however, the underreporting of sexual violence experiences is likely to remain an issue.

Second, there are limitations of the questions asked. For instance, participants were assessed about sexual violence experiences in the previous 12 months, meaning that experiences before this period might not have been reported. The questions about sexual violence were dichotomous, hence it was not possible to investigate important aspects such as severity, frequency, and perpetrator. Lower severity experiences such as harassment but experienced constantly might have substantial impacts. The sexual violence measures included also do not include the full range of these potential experiences, for instance online harassment or experiences over text were not asked about and young people might not have reported these the unwelcome sexual approach question; better and more specific questions are needed to fully understand the full impact of sexual violence at this age.

The lack of data on these aspects is a limitation, and more studies need to collect this information from adolescents. Third, potential unmeasured confounders (e.g., parental neglect, violence between parents) not captured in this study might attenuate the observed effects. Despite PSM being a very useful technique to be used in observational studies where RCTs are not possible, while matching can improve efficiency and precision, it is limited in reducing confounding by unobserved variables. Future research should aim to triangulate these findings using alternative approaches to assess the impact of sexual violence on mental health.

As previously highlighted, most research and policy focus for sexual violence is directed to either child sexual abuse or interpersonal violence within relationships in older adolescents and adults, however, a third category of violence which is experienced in community settings on the streets, by peers, in schools, and online, is also relevant and especially salient as children transition into adolescence and adulthood. The lack of population-based datasets that include detailed measures on the perpetrators, frequency, severity, types, and settings for sexual violence severely restricts the capacity of research and policy to estimate the real consequences of these different experiences, which would help target appropriate policy and support. This stresses the importance of formulating more comprehensive research questions about sexual violence experiences as well as the need to gather more data on such experiences in adolescence. Importantly, sexual violence is known to be widespread in adulthood as well,^32^ thus suggesting a continuation and potential worsening of mental health sequalae of these experiences. Because sexual violence is a global phenomenon, it is imperative to increase efforts to understand its role in the mental health of adolescent girls while also promoting the development of effective preventative strategies, both to prevent sexual violence and to mitigate their subsequent impacts when it does occur.

The implications of our findings are clear and multifold. A reduction in sexual violence is likely to lead to a decrease in mental health problems, this can in turn reduce the societal costs not only for the individuals affected, but also for their families and wider society.^33^ Starting early, interventions aimed at educating young people in schools with regards to sexual violence are one actionable avenue.^34^ Moreover, more attention should be paid at the societal level by policy makers on reducing the general societal tolerance and permissiveness currently shown for sexual violence.^35^ The low conviction rates for perpetrators have severe impacts on victims’ mental health,^36^ and erode trust in the law enforcement systems that are meant to protect victims. Legal and law enforcement changes might reduce risks, for example by tackling the existing policies regarding prosecutions and more sexual violence deterrents. Due to the traumatic nature of these experiences, acknowledging this in the responses and support offered to victims is important,^37^ as well as reducing victim-blaming that is prevalent in the law enforcement and legal systems.^38^ The effect sizes observed also highlight the urgent need for better tailored and targeted support for victims, to try and mitigate the long-term mental health impacts.

## Conclusion

This study aimed to estimate the impact of sexual violence experienced in mid-adolescence on mental health difficulties using a robust methodology in a longitudinal population-based cohort study. The findings highlight the large burden of sexual violence experienced at this age and stress the importance of reducing these experiences, especially for girls. Based on our findings, if sexual violence experiences in adolescence could be eliminated, 14.0-18.7% fewer teenage girls might engage in self-harming and attempt suicide in mid-adolescence. Addressing this risk factor would also contribute substantially to closing the observed gender gap in these mental health outcomes that emerge in adolescence. The findings of this study emphasise the need to consider this common and important, yet often gendered and understudied, risk factor more seriously and urgently in adolescent mental health research, clinical practice, and policymaking.

## Data Availability

The Millennium Cohort Study data is available to use by researchers via the UK Data Service (https://ukdataservice.ac.uk/).

## Acknowledgements

This study was supported by grant MR/N013867/1 from the Medical Research Council. The Millennium Cohort Study is supported by the Economic and Social Research Council and a consortium of UK government departments. The funders of the study had no role in study design, data collection, data analysis, data interpretation, or writing of this report.

The authors are grateful for the cooperation of the Millennium Cohort Study families who voluntarily participate in the study. They would also like to thank a large number of stakeholders from academic, policymaker, and funder communities and colleagues at the Centre for Longitudinal Studies involved in data collection and management. We are also grateful for the feedback provided on an earlier draft of the manuscript by Drs. Francesca Solmi and Gemma Lewis, Division of Psychiatry, UCL.

## Declaration of Interest

FB and PP declare no conflicts of interest.

## Contributions

PP conceptualised, designed and supervised the study. FB led on literature review, data management and analysis, with PP verifying analyses. Both authors drafted and revised the manuscript, generated the visualisations, had access to the data, and approved submission of the manuscript for publication.

## Notes

### Competing Interest Statement

The authors have declared no competing interest.

### Funding Statement

This study was supported by grants MR/N013867/1 from the Medical Research Council. The Millennium Cohort Study is supported by the Economic and Social Research Council and a consortium of UK government departments. The funders of the study had no role in study design, data collection, data analysis, data interpretation, or writing of this report.

### Summary of Updates

We updated the estimates for both girls and boys after having re-checked the code for the analytic sample and having found a mistake in the syntax, which has now been fixed. The estimates have been updated throughout, including manuscript, tables, figures, and supplementary material.

## References

1. van de Velde S, Bracke P, Levecque K. Gender differences in depression in 23 European countries. Cross-national variation in the gender gap in depression. Social Science & Medicine [Internet]. 2010;71(2):305–13. Available from: https://www.sciencedirect.com/science/article/pii/S0277953610002844

2. Patalay P, Fitzsimons E. Psychological distress, self-harm and attempted suicide in UK 17-year olds: prevalence and sociodemographic inequalities. The British Journal of Psychiatry. 2021;1–3.

3. Piccinelli M, Wilkinson G. Gender differences in depression: Critical review. The British Journal of Psychiatry. 2000;177(6):486–92.

4. Hyde JS, Mezulis AH, Abramson LY. The ABCs of depression: integrating affective, biological, and cognitive models to explain the emergence of the gender difference in depression. Psychol Rev. 2008;115(2):291.

5. Eom E, Restaino S, Perkins AM, Neveln N, Harrington JW. Sexual harassment in middle and high school children and effects on physical and mental health. Clinical Pediatrics. 2015;54(5):430–8.

6. Finkelhor D, Shattuck A, Turner HA, Hamby SL. The lifetime prevalence of child sexual abuse and sexual assault assessed in late adolescence. Journal of adolescent Health. 2014;55(3):329–33.

7. Elkin M. The lasting impact of violence against women and girls. Office for National Statistics. 2021.

8. Karatekin C. Adverse childhood experiences (ACEs), stress and mental health in college students. Stress and Health. 2018;34(1):36–45.

9. Schilling EA, Aseltine RH, Gore S. Adverse childhood experiences and mental health in young adults: a longitudinal survey. BMC Public Health. 2007;7(1):1–10.

10. Carey KB, Norris AL, Durney SE, Shepardson RL, Carey MP. Mental health consequences of sexual assault among first-year college women. Journal of American college health. 2018;66(6):480–6.

11. Rothman K, Georgia Salivar E, Roddy MK, Hatch SG, Doss BD. Sexual assault among women in college: Immediate and long-term associations with mental health, psychosocial functioning, and romantic relationships. J Interpers Violence. 2021;36(19–20):9600–22.

12. Herbert A, Heron J, Barter C, Szilassy E, Barnes M, Howe LD, et al. Risk factors for intimate partner violence and abuse among adolescents and young adults: findings from a UK population-based cohort. Wellcome Open Res. 2020;5.

13. Khadr S, Clarke V, Wellings K, Villalta L, Goddard A, Welch J, et al. Mental and sexual health outcomes following sexual assault in adolescents: a prospective cohort study. The Lancet Child & Adolescent Health. 2018;2(9):654–65.

14. Baiden P, Panisch LS, Kim YJ, LaBrenz CA, Kim Y, Onyeaka HK. Association between first sexual intercourse and sexual violence victimization, symptoms of depression, and suicidal behaviors among adolescents in the United States: findings from 2017 and 2019 National Youth Risk Behavior Survey. Int J Environ Res Public Health. 2021;18(15):7922.

15. Landstedt E, Gillander K, Din GÅ. Deliberate self-harm and associated factors in 17-year-old Swedish students. Scandinavian Journal of Public Health. 2011;39:17–25.

16. Bendixen M, Daveronis J, Kennair LEO. The effects of non-physical peer sexual harassment on high school students’ psychological well-being in Norway: consistent and stable findings across studies. International Journal of Public Health. 2018;63(1):3–11.

17. Rinehart SJ, Espelage DL, Bub KL. Longitudinal Effects of Gendered Harassment Perpetration and Victimization on Mental Health Outcomes in Adolescence. Journal of Interpersonal Violence. 2017 Aug 21;35(23–24):5997–6016.

18. Chiodo D, Wolfe DA, Crooks C, Hughes R, Jaffe P. Impact of Sexual Harassment Victimization by Peers on Subsequent Adolescent Victimization and Adjustment: A Longitudinal Study. Journal of Adolescent Health. 2009 Sep 1;45(3):246–52.

19. Dahlqvist HZ, Landstedt E, Young R, Gådin KG. Dimensions of Peer Sexual Harassment Victimization and Depressive Symptoms in Adolescence: A Longitudinal Cross-Lagged Study in a Swedish Sample. Journal of Youth and Adolescence. 2016;45(5):858–73.

20. Marshall SK, Faaborg-Andersen P, Tilton-Weaver LC, Stattin H. Peer sexual harassment and deliberate self-injury: Longitudinal cross-lag investigations in Canada and Sweden. Journal of Adolescent Health. 2013;53(6):717–22.

21. Rosenbaum PR, Rubin DB. The central role of the propensity score in observational studies for causal effects. Biometrika. 1983;70(1):41–55.

22. Joshi H, Fitzsimons E. The Millennium Cohort Study: the making of a multi-purpose resource for social science and policy. Longitudinal and Life Course Studies. 2016;7(4):409–30.

23. Kessler RC, Barker PR, Colpe LJ, Epstein JF, Gfroerer JC, Hiripi E, et al. Screening for serious mental illness in the general population. Arch Gen Psychiatry. 2003;60(2):184–9.

24. Leuven E, Sianesi B. PSMATCH2: Stata module to perform full Mahalanobis and propensity score matching, common support graphing, and covariate imbalance testing. 2003;

25. Mansournia MA, Altman DG. Population attributable fraction. BMJ (Online). 2018;360(February):2–3.

26. Newson R. PUNAF: Stata module to compute population attributable fractions for cohort studies. 2010;

27. Ofsted. Review of sexual abuse in schools and colleges. 2021;

28. Miller E. Prevention of and Interventions for Dating and Sexual Violence in Adolescence. Pediatric Clinics of North America. 2017;64(2):423–34.

29. Kennedy AC, Prock KA. “I still feel like I am not normal”: A review of the role of stigma and stigmatization among female survivors of child sexual abuse, sexual assault, and intimate partner violence. Trauma, Violence, & Abuse. 2018;19(5):512–27.

30. Lundgren R, Amin A. Addressing intimate partner violence and sexual violence among adolescents: emerging evidence of effectiveness. Journal of Adolescent Health. 2015;56(1):S42–50.

31. Langeland W, Smit JH, Merckelbach H, de Vries G, Hoogendoorn AW, Draijer N. Inconsistent retrospective self-reports of childhood sexual abuse and their correlates in the general population. Soc Psychiatry Psychiatr Epidemiol. 2015;50(4):603–12.

32. de Toledo Blake M, Drezett J, Vertamatti MA, Adami F, Valenti VE, Paiva AC, et al. Characteristics of sexual violence against adolescent girls and adult women. BMC Womens Health. 2014;14(1):1–7.

33. Trautmann S, Rehm J, Wittchen HU. The economic costs of mental disorders: Do our societies react appropriately to the burden of mental disorders? EMBO Rep. 2016/08/04. 2016 Sep;17(9):1245–9.

34. Letourneau EJ, Schaeffer CM, Bradshaw CP, Feder KA. Preventing the Onset of Child Sexual Abuse by Targeting Young Adolescents With Universal Prevention Programming. Child Maltreat. 2017/01/01. 2017 May;22(2):100–11.

35. Brown CS, Biefeld SD, Elpers N. A Bioecological Theory of Sexual Harassment of Girls: Research Synthesis and Proposed Model. Review of General Psychology. 2020 Sep 10;24(4):299–320.

36. Bohner G, Eyssel F, Pina A, Siebler F, Viki GT. Rape myth acceptance: Cognitive, affective and behavioural effects of beliefs that blame the victim and exonerate the perpetrator. In: Rape. Willan; 2013. p. 40–68.

37. Harper DJ, Cromby J. From ‘what’s wrong with you?’To ‘what’s happened to you?’: an introduction to the special issue on the power threat meaning framework. Journal of Constructivist Psychology. 2022;35(1):1–6.

38. Greeson MR, Campbell R, Fehler-Cabral G. “NOBODY DESERVES THIS”: ADOLESCENT SEXUAL ASSAULT VICTIMS’PERCEPTIONS OF DISBELIEF AND VICTIM BLAME FROM POLICE. Journal of Community Psychology. 2016;44(1):90–110.

