## Supplementary material for "The impact of sexual violence in mid-adolescence on mental health: a UK population-based longitudinal study"

**Figure S1. Study design**

Directed Acyclical Graph (DAG) informing the multivariable regression

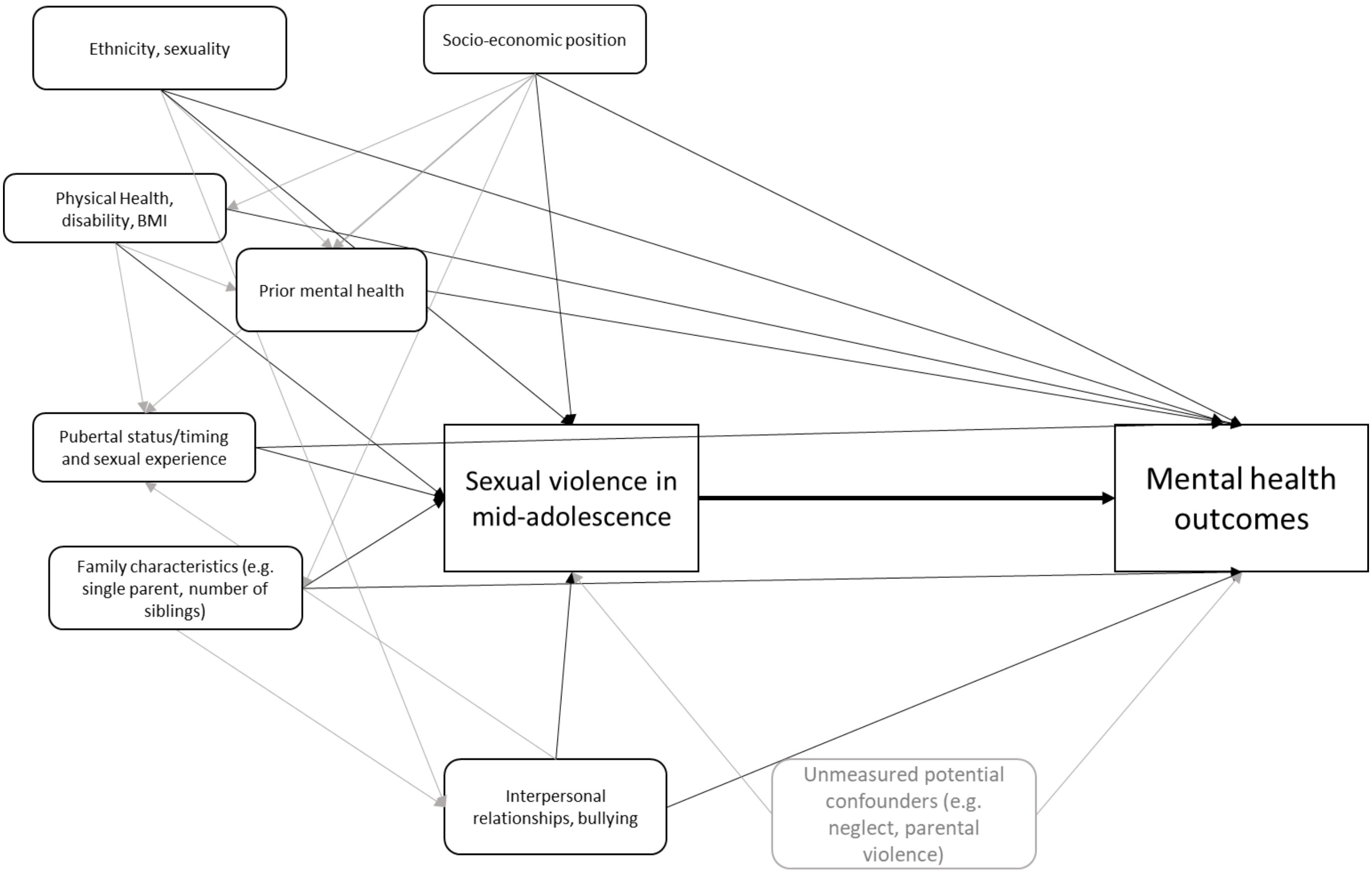

Design for the matched pseudo-experimental approach

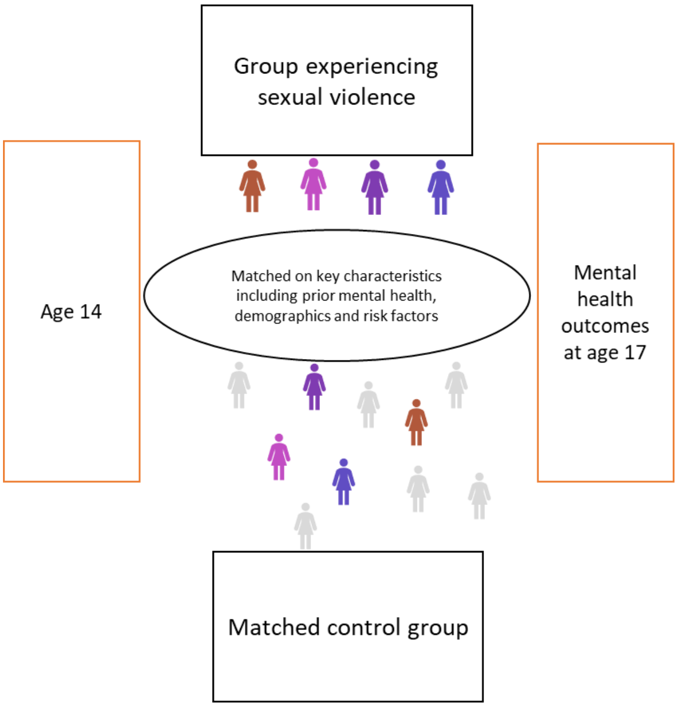

In this matching approach, each individual from the treatment group is matched to an individual in the control group based on similarity on pre-specified criteria.

The nature of sexual violence at this age, which is not an experience that an individual selects into, but rather is done to them, also increases the methodological scope for inferring causality using such pseudo-experimental approaches (in contrast to smoking or physical activity, for instance, which often involve self-selection into these behaviours).

The quality of these analyses rests on the assumption that a counterfactual group can be identified who are similar in other key ways except for their experiences of sexual violence.

### Rationale and literature for confounders included

We chose the following matching and confounding factors in line with previous literature indicating the following as being potentially relevant confounders based on prior evidence and theoretical rationale.

Prior mental health was controlled for as it allowed a focus on mental health as developed in mid-adolescence and is a risk factor for experiencing sexual violence [1]. Depressive symptoms at age 14 was assessed using the Short Mood and Feelings Questionnaire. We also controlled for self-harm at age 14 as a prior measure of self-harm until 14 years [2].

Socio-demographic characteristics were included as potential confounders as they predict both risk of sexual violence and mental health outcomes. These included Ethnicity, as being part of a minority is a risk factor for sexual assault [3], and there are established ethnic differences for poor mental health at this age [4]. Socio-economic factors were included as socio-economic disadvantage is expected to increase the risk of both sexual violence and mental health difficulties. Various different dimensions of socio-economic status were included including parental education [1] and family income [1]. Sexual identity was included as sexual minorities experience higher levels of assault, violence, and mental health difficulties [3,5]. Household composition and family structure variables were included as risk factors for both sexual violence and mental health, including whether it was a single parent household [1,6], number of siblings [7,8].

We controlled for puberty-related characteristics as early pubertal development [9] and menarche (in girls) [10] has been linked to early sexual activity, greater risk taking and differences in mental health outcomes. Early sexual activity [10] was also included as it might increase the likelihood of experience sexual violence. We also control for prior sexual violence prior to age 14, and this will include child sexual abuse and might be an important confounder that reflects several other early childhood risk factors.

We further adjusted the analyses for interpersonal relationships, including poor peer relationships [10] and relationship status [11], due to their association with sexual violence; and having been bullied [12,13], which has been shown to be associated with both sexual harassment and with poor mental health outcomes.

Finally, we included health and risk-related variables. Health-related characteristics included body mass index (BMI), as it has been shown to be longitudinally associated with mental health and risk of bullying and assault [14]. Risk-related variables were risky behaviours [14,15] such as smoking cigarettes, consuming alcohol, and taking drugs. We additionally controlled for whether the participants missed school without parents' permission, as this might increase the risk of being in situations that are less safe [16]. Finally, we included other dimensions of health and wellbeing such as life satisfaction [17] and disability/chronic illness/special needs [1,18], as evidence shows that they are associated with sexual violence and mental health outcomes.

### Measures

Girls and boys: We stratify all analysis. In MCS information on biological sex is available for all participants and at age 17 cohort members also reported on their gender identity. When we started this project we a priori focussed on girls, and included all individuals whose sex was female and identified as female. We included boys based on peer reviewer feedback and at that stage, to avoid individuals being in both samples, boys in this study are cohort members whose sex is male and who did not identify as female.

**Choice of Psychological distress measure:** The Kessler-6 (K-6) [19] is a self-administered measure of psychological distress which includes six items about depressive and anxiety symptoms. Specifically, individuals are asked "During the last 30 days, about how often did you feel 1) so depressed that nothing could cheer you up; 2) hopeless; 3) restless or fidgety; 4) that everything was an effort; 5) worthless; 6) nervous?". Responses are scored on a 5-point Likert scale: 4 = all of the time; 3 = most of the time; 2 = some of the time; 1 = a little of the time; 0 = none of the time. The minimum score is 0 and the maximum score is 24, and the higher the score the higher the level of psychological distress experienced by the individual.

The measure was selected as it is the main measure capturing symptoms of common mental health conditions in the MCS cohort. The cohort include this measure as it is a well-validated short measure of population psychological distress with established clinical thresholds based on diagnosis to capture clinical levels of symptoms. The measure is used widely, especially for epidemiological research in a wide range of countries and languages including in Swahili, [20] Korean,[21] Chinese, [21] [22] Japanese, [23] Arabic, [24] and Vietnamese.[25] Studies comparing the measure to other widely used measures such as PHQ and GAD have found it to be a useful screener and measure of treatment progress in clinical settings as well.[26]

The measure is free to use and does not require payment.

### Details of confounding and matching variables used

Depressive symptoms were assessed at 14 years, however, we used the Short Mood and Feelings Questionnaire (SMFQ; [27]) rather than the K-6 because, this was the measure used to assess adolescent symptoms at age 14. The SMFQ had been previously validated [28] and is a 13-item self-report questionnaire with a 3-point Likert scale (not true = 0, sometimes true = 1, true = 2) and a total score ranging from 0 to 26, and we used 12 as cut-off. Items include feelings of unhappiness, crying a lot, etc., and higher scores indicate higher levels of depressive symptoms. Self-harm at age 14 was assessed with a single question on whether participants self-harmed or not in the past year.

Information on participants' ethnicity based on parent reports was used to distinguish among White, Mixed, Indian, Pakistani and Bangladeshi, Black or Black British, and other ethnic groups. Socioeconomic status (SES) was indicated by parental education measured with the National Vocational Qualification (NVQ) and was treated as a binary variable indicating the presence or not of a higher degree. Family income was categorised according to the Organisation for Economic Co-operation and Development (OECD) equivalised income quintiles (from 1 to 5), based upon UK income distribution. We also included number of siblings (total number), including natural, half, step, adopted and foster siblings; and single parent household, indicating the number (one or two) of parents in one household. Sexual identity at age 17 was explored and recoded to indicate whether participants thought about themselves as heterosexual or sexual minority.

Pubertal status at age 14 (Not yet started to grow, Started to grow) was measured by assessing body hair, breast growth, and begin of menstruation for females, and by assessing body hair, voice change, and facial hair in males, and the presence of any of these physical signs indicated the start of puberty. For age of menarche (just females; Before 11 years, At or after 11 years) we used age 11 years as a cut-off to distinguish girls who experienced menstruation prior to that age from girls who began menses at that age or later. We chose age 11 because the NHS states that this is the average age for girls to start puberty (https://www.nhs.uk/conditions/early-or-delayed-puberty/).

Sexual violence prior to age 14 was recorded using a single question from the 'Victimisation grid': "In the past 12 months has anyone made an unwelcome sexual approach to you or assaulted you sexually?" (no = 0, yes = 1).

Single items related to early sexual relationship (No early sexual activity, Early sexual activity: put/being put hands under clothes, touched/being touched private parts, performed/being performed oral sex, having had sexual intercourse), relationship status (Doesn't have a boyfriend/girlfriend, Has a boyfriend/girlfriend) and peer relationships (Doesn't have any close friends, Has close friends) were also included in the analyses. Three items about the frequency of experiencing of being bullied (how often brothers/sisters hurt or pick on them, how often other children bullied them online, how often other children hurt or pick on them), were recoded and combined to create a categorical variable on how often participants were bullied (never = 0, sometimes = 1, often = 2).

Body mass index (BMI) was derived from objectively assessed height and weight. Missing school without parents' permission (missed school: no/yes à how often) was recoded and combined to create a categorical variable on how often participants missed school (never = 0, sometimes = 1, often = 2). Risky behaviours included items on whether participants ever smoked, consumed alcohol, or took drugs (including cannabis); moreover, items related to alcohol consumption were recoded and combined to create a variable on how often participants had five or more alcoholic drink at a time (never = 0, sometimes = 1, often = 2, very often = 3). Lifelong disability was measured combining three items on disability, chronic illnesses and special needs (has a statement of special needs, has longstanding illness, their conditions/illnesses limit everyday activities), and was treated as a single binary variable (yes/no). Life satisfaction was assessed using a 'Wellbeing grid' including six items (how happy they are with family, friends, life as a whole, school, school work, the way they look) related to participants' general satisfaction with family, friends, life as a whole, school, school work, and the way they look; these items were then recoded so that rating ranged from 0 ('Not happy at all') to 6 ('Completely happy'), and a total score was derived with the maximum possible score being 36.

Matching variables: depressive symptoms at age 14, self-harm at age 14, sexual violence reported at age 14, ethnicity, parental education, pubertal status, sexual identity.

Confounding variables: family income, number of siblings, single parent household, age of menarche, early sexual activity, relationship status, peer relationship quality, bullying, BMI, missing school without parents' permission, risky behaviours (smoking, drinking alcohol, taking drugs), life satisfaction, lifelong disability/illness.

### Ethics Statement

Ethical approval for each sweep of the Millennium Cohort Study was received at the point of the sweep from National Research Ethics Committees.

Ethical approval for age 17 sweep was received from the National Research Ethics

Service (NRES) Research Ethics Committee (REC) North East – York (REC ref: **17/NE/0341).**

Ethical approval for all the other sweeps:

Sweep 1 (9 months): South West MREC (MREC/01/6/19)

Sweep 2 (3 years): London MREC (MREC/03/2/022)

Sweep 3 (5 years): London MREC Committee (05/MRE02/46)

Sweep 4 (7 years): Yorkshire MREC (07/MRE03/32)

Sweep 5 (11 years): Yorkshire and The Humbler – Leeds East (11/YH/0203)

Sweep 6 (14 years): London – Central MREC (13/LO/1786)

There was a detailed protocol in place for both confidentiality and aspects of respondent wellbeing. These are detailed in the MCS technical report for this sweep [29], and included resources such as information about professional sources of help and a helpline number.

### Descriptives for each sexual violence measure

#### Girls

**Unwelcomed sexual approach**
Sexual approach yes: 991/5,119 = 19.4%
Sexual approach no: 4,120/5,119 = 80.6%
Missing data: 8/5,119 = 0.2%

**Sexual assault**
Sexual assault yes: 269/5,119 = 5.3%
Sexual assault no: 4,847/5,119 = 94.7%
Missing data: 3/5,119 = 0.1%

**Any sexual violence**
Any sexual violence yes: 1,035/5,119 = 20.2%
Any sexual violence no: 4,084/5,119 = 79.8%

#### Boys

**Unwelcomed sexual approach**
Sexual approach yes: 251/4,852 = 5.2%
Sexual approach no: 4,601/4,852 = 94.8%

**Sexual assault**
Sexual assault yes: 50/4,850 = 1.0%
Sexual assault no: 4,734/4,782 = 98.9%
Missing data: 2/4,782 = 0.04%

**Any sexual violence**
Any sexual violence yes: 263/4,852 = 5.4%
Any sexual violence no: 4,589/4,852 = 94.6%

**Figure S2. Venn diagrams showing overlaps of measures in females and males**

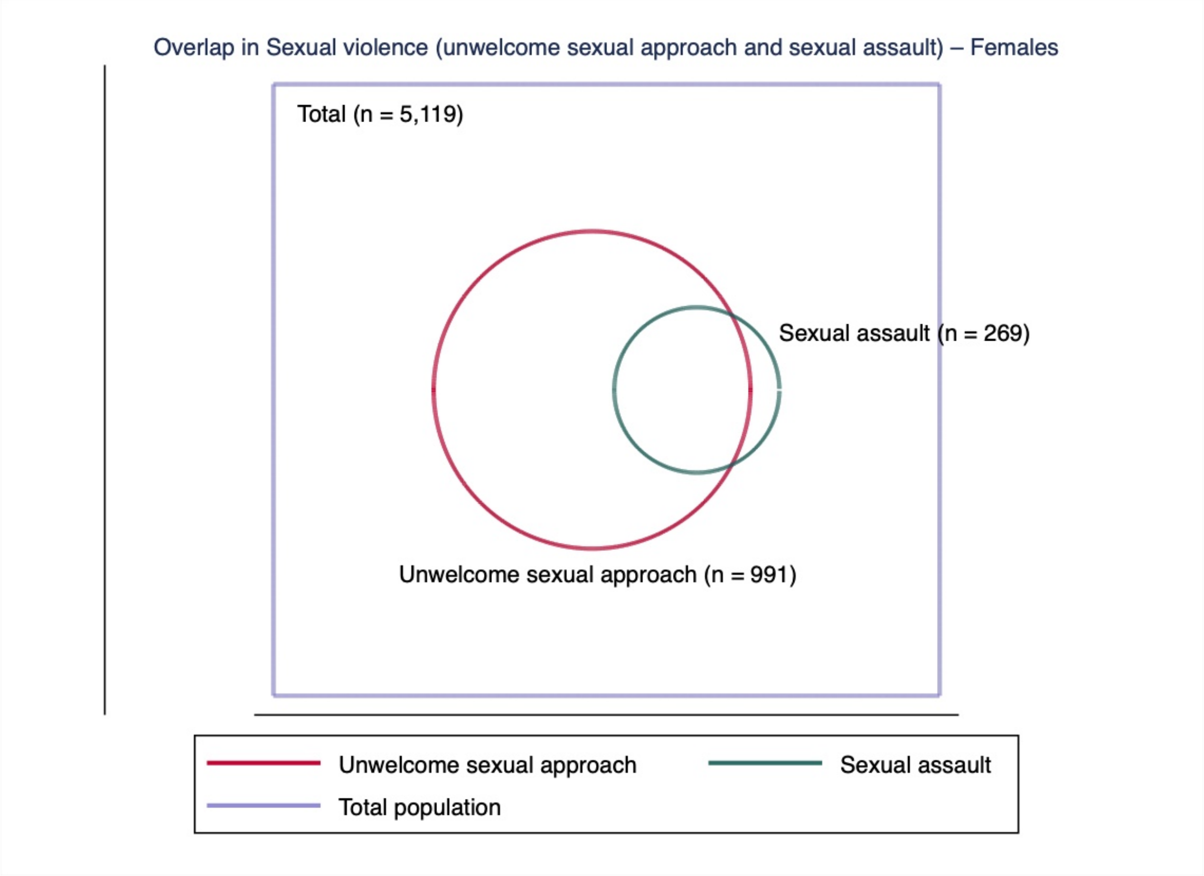

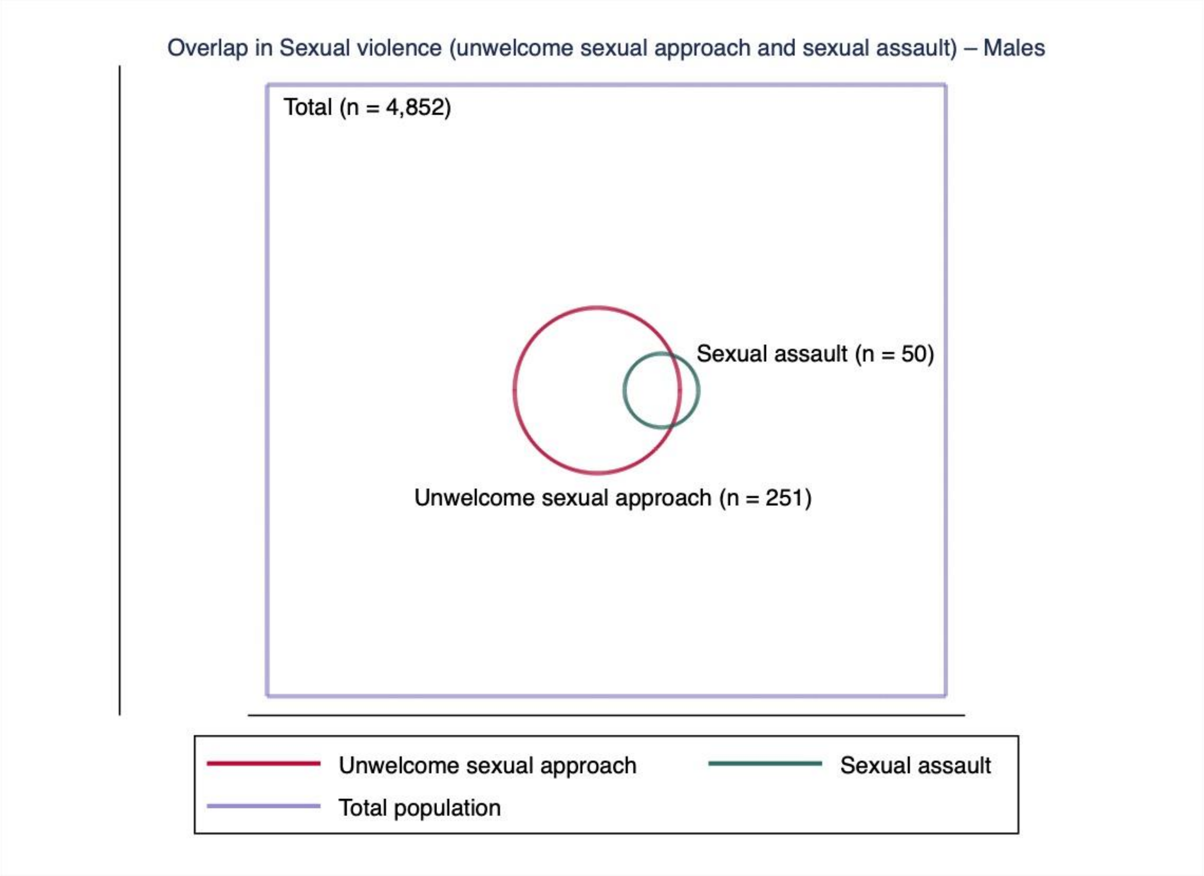

### Table S1. Descriptive analysis of mental health outcomes in the PSM samples

| | Girls | | | Boys | | |
|---|---|---|---|---|---|---|
| | **PSM sample (N = 4,547)** | **PSM: Sexual violence group (N = 938)** | **PSM: Matched control group (N = 938)** | **PSM sample (N = 4,235)** | **PSM: Sexual violence group (N = 230)** | **PSM: Matched control group (N = 230)** |
| | **N (%) or M (SE) – [95% CI]** | **N (%) or M (SE) – [95% CI]** | **N (%) or M (SE) – [95% CI]** | **N (%) or M (SE) – [95% CI]** | **N (%) or M (SE) – [95% CI]** | **N (%) or M (SE) – [95% CI]** |
| **Mental health at age 17** | | | | | | |
| ***Psychological distress (continuous)*** | | | | | | |
| | 10.01 (0.12) – [9.77, 10.24] | 10.66 (0.16) – [10.34, 10.98] | 9.36 (0.17) – [9.02, 9.70] | 8.06 (0.24) – [7.60, 8.53] | 9.43 (0.34) – [8.77, 10.09] | 6.70 (0.31) – [6.09, 7.30] |
| ***Psychological distress (binary)*** | | | | | | |
| < 13 tot score | 1,276 (68.1) – [65.9, 70.2] | 595 (63.5) – [60.4, 66.5] | 681 (72.7) – [69.7, 75.4] | 371 (80.7) – [76.8, 84.0] | 171 (74.3) – [68.3, 79.6] | 200 (87.0) – [81.9, 90.7] |
| ≥ 13 tot score | 598 (31.9) – [29.8, 34.1] | 342 (36.5) – [33.5, 39.6] | 256 (27.3) – [24.6, 30.3] | 89 (19.3) – [16.0, 23.2] | 59 (25.7) – [20.4, 31.7] | 30 (13.0) – [9.3, 18.1] |
| ***Self-harm*** | | | | | | |
| No | 1,041 (56.6) – [54.3, 58.8] | 445 (48.7) – [45.5, 52.0] | 596 (64.3) – [61.1, 67.3] | 296 (65.2) – [60.7, 69.5] | 121 (54.0) – [47.4, 60.5] | 175 (76.1) – [70.1, 81.2] |
| Yes | 799 (43.4) – [41.2, 45.7] | 468 (51.3) – [48.0, 54.5] | 331 (35.7) – [32.7, 38.9] | 158 (34.8) – [30.5, 39.3] | 103 (46.0) – [39.5, 52.6] | 55 (23.9) – [18.8, 29.9] |
| ***Attempted suicide*** | | | | | | |
| No | 1,538 (83.9) – [82.1, 85.5] | 745 (81.7) – [79.0, 84.1] | 793 (86.0) – [83.6, 88.1] | 400 (87.9) – [84.6, 90.6] | 187 (83.1) – [77.6, 87.5] | 213 (92.6) – [88.4, 95.4] |
| Yes | 296 (16.1) – [14.5, 17.9] | 167 (18.3) – [15.9, 21.0] | 129 (14.0) – [11.9, 16.4] | 55 (12.1) – [9.4, 15.4] | 38 (16.9) – [12.5, 22.4] | 17 (7.4) – [4.6, 11.6] |
| **Mental health at age 14** | | | | | | |
| ***Psychological distress (continuous)*** | | | | | | |
| | 9.69 (16.72) – [9.37, 10.02] | 9.57 (0.23) – [9.11, 10.03] | 9.82 (0.24) – [9.35, 10.29] | 6.24 (0.28) – [5.68, 6.79] | 6.06 (0.38) – [5.30, 6.81] | 6.42 (0.42) – [5.59, 7.24] |
| ***Psychological distress (binary)*** | | | | | | |
| < 12 tot score | 1,181 (63.0) – [60.7, 65.1] | 600 (64.0) – [60.8, 67.0] | 581 (61.9) – [58.8, 65.0] | 378 (82.2) – [78.4, 85.4] | 192 (83.5) – [78.1, 87.8] | 186 (80.9) – [75.2, 85.5] |
| ≥ 12 tot score | 695 (37.0) – [34.9, 39.3] | 338 (36.0) – [33.0, 39.2] | 357 (38.1) – [35.0, 41.2] | 82 (17.8) – [14.6, 21.6] | 38 (16.5) – [12.2, 21.9] | 44 (19.1) – [14.5, 24.8] |
| ***Self-harm*** | | | | | | |
| No | 1,195 (63.7) – [61.5, 65.8] | 598 (63.8) – [60.6, 66.8] | 597 (63.6) – [60.5, 66.7] | 373 (81.1) – [77.2, 84.4] | 187 (81.3) – [75.7, 85.9] | 186 (80.9) – [75.2, 85.5] |
| Yes | 681 (36.3) – [34.2, 38.5] | 340 (36.3) – [33.2, 39.4] | 341 (36.4) – [33.3, 39.5] | 87 (18.9) – [15.6, 22.8] | 43 (18.7) – [14.1, 24.3] | 44 (19.1) – [14.5, 24.8] |

PSM = propensity score matching.

**Table S2. Regression coefficients and risk ratios for the association between sexual violence and mental health in non-imputed data (complete case analysis)**

| | Psychological distress (continuous) | Psychological distress (binary) | Self-harm | Attempted Suicide |
|---|---|---|---|---|
| | **N**<br>**Coefficient [95% CI]** | **N**<br>**RR [95% CI]** | **N**<br>**RR [95% CI]** | **N**<br>**RR [95% CI]** |
| **Girls** | | | | |
| Unadjusted | 5,115 | 5,115 | 4,956 | 4,946 |
| | 3.00 [2.66, 3.35] | 0.72 [0.62, 0.82] | 0.79 [0.71, 0.87] | 0.86 [0.70, 1.03] |
| Adjustment 1 | 4,613 | 4,613 | 4,531 | 4,521 |
| | 1.71 [1.38, 2.04] | 0.39 [0.28, 0.50] | 0.52 [0.43, 0.60] | 0.38 [0.21, 0.56] |
| Adjustment 2 | 4,583 | 4,583 | 4,502 | 4,492 |
| | 1.60 [1.28, 1.93] | 0.38 [0.27, 0.49] | 0.46 (0.38, 0.55] | 0.39 [0.21, 0.57] |
| Adjustment 3 | 3,974 | 3,974 | 3,902 | 3,895 |
| | 1.72 [1.37, 2.07] | 0.41 [0.29, 0.53] | 0.47 [0.37, 0.56] | 0.36 [0.17, 0.55] |
| Adjustment 4 | 3,973 | 3,973 | 3,901 | 3,894 |
| | 1.67 [1.32, 2.01] | 0.39 [0.28, 0.51] | 0.46 (0.36, 0.55] | 0.36 [0.16, 0.55] |
| Adjustment 5 | 3,758 | 3,758 | 3,692 | 3,686 |
| | 1.63 [1.28, 1.99] | 0.40 [0.28, 0.52] | 0.44 [0.34, 0.54] | 0.31 [0.11, 0.51] |
| **Boys** | | | | |
| Unadjusted | 4,850 | 4,850 | 4,758 | 4,760 |
| | 3.65 [3.11, 4.19] | 1.12 [0.90, 1.34] | 1.08 [0.93, 1.23] | 1.57 [1.26, 1.87] |
| Adjustment 1 | 4,317 | 4,317 | 4,266 | 4,268 |
| | 2.62 [2.10, 3.14] | 0.78 [0.54, 1.03] | 0.84 [0.67, 1.00] | 1.14 [0.80, 1.48] |
| Adjustment 2 | 4,294 | 4,294 | 4,242 | 4,244 |
| | 2.34 [1.82, 2.87] | 0.70 [0.44, 0.96] | 0.76 [0.58, 0.93] | 1.10 [0.74, 1.47] |
| Adjustment 3 | 4,181 | 4,181 | 4,132 | 4,133 |
| | 2.32 [1.79, 2.86] | 0.69 [0.42, 0.97] | 0.73 [0.55, 0.91] | 1.03 [0.66, 1.41] |
| Adjustment 4 | 4,172 | 4,172 | 4,123 | 4,124 |
| | 2.30 [1.77, 2.83] | 0.70 [0.43, 0.97] | 0.71 [0.52, 0.89] | 1.08 [0.70, 1.46] |
| Adjustment 5 | 4,051 | 4,051 | 4,003 | 4,004 |
| | 2.21 [1.67, 2.75] | 0.64 [0.35, 0.92] | 0.68 [0.50, 0.87] | 1.04 [0.64, 1.45] |

RR = risk ratio; CI = confidence interval.

**Table S3a. Descriptive analyses – Exposure & outcomes by sample (girls)**

| Exposure & outcomes | PSM sample | | Full analytic sample | |
|---|---|---|---|---|
| | **Sample** | **Missingness** | **Sample** | **Missingness** |
| | **N (%)** | **N (%)** | **N (%)** | **N (%)** |
| **Total obs.** | **4,547 (100.0)** | | **5,119 (100.0)** | |
| **Sexual violence <17 years** | 4,547 (100.0) | 0 (0) | 5,119 (100.0)* | 0 (0) |
| No | 3,609 (79.4) | | 4,084 (79.8)** | |
| Yes | 938 (20.6) | | 1,035 (20.2)** | |
| **Sexual violence <14 years** | 4,551 (100.0) | 0 (0) | 4,661 (91.1) | 458 (8.9) |
| No | 4,352 (95.6) | | 4,462 (95.7) | |
| Yes | 199 (4.4) | | 199 (4.3) | |
| **Psychological distress 17 years** | 4,548 (99.9) | 3 (0.07) | 5,115 (99.9) | 4 (0.08) |
| < 13 tot score | 3,546 (78.0) | | 3,974 (77.7) | |
| ≥ 13 tot score | 1,002 (22.0) | | 1,141 (22.3) | |
| **Self-harm 17 years** | 4,468 (98.2) | 83 (1.8) | 4,956 (96.8) | 163 (3.2) |
| No | 3,166 (70.9) | | 3,520 (71.0) | |
| Yes | 1,302 (29.1) | | 1,436 (29.0) | |
| **Suicide attempt 17 years** | 4,459 (98.0) | 92 (2.0) | 4,946 (96.6) | 173 (3.4) |
| No | 4,020 (90.2) | | 4,437 (89.7) | |
| Yes | 439 (9.8) | | 509 (10.3) | |
| | **Mean (SD)** | | **Mean (SD)** | |
| **Psychological distress 17 years** | 8.38 (5.12) | 3 (0.07) | 8.36 (5.14) | 4 (0.08) |

The following rules apply to all the variables in the table:
* % uses whole sample as denominator
** % uses sample with complete data on a specific variable as denominator
N = number of participants; % = percentage of the number of participants; SD = standard deviation; PSM = propensity score matching; MI = multiple imputation.

### Table S3b. Descriptive analyses – Exposure & outcomes by sample (boys)

| Exposure & outcomes | PSM sample | | Full analytic sample | |
|---|---|---|---|---|
| | Sample N (%) | Missingness N (%) | Sample N (%) | Missingness N (%) |
| **Total obs.** | **4,235 (100.0)** | | **4,852 (100.0)** | |
| **Sexual violence <17 years** | 4,235 (100.0) | 0 (0) | 4,852 (100.0)* | 0 (0) |
| No | 4,005 (94.6) | | 4,589 (94.6)** | |
| Yes | 230 (5.4) | | 263 (5.4)** | |
| **Sexual violence <14 years** | 4,235 (100.0) | 0 (0) | 4,344 (89.5) | 508 (10.5) |
| No | 4,184 (98.8) | | 4,289 (98.7) | |
| Yes | 51 (1.2) | | 55 (1.3) | |
| **Psychological distress 17 years** | 4,234 (99.9) | 1 (0.02) | 4,850 (99.9) | 2 (0.04) |
| < 13 tot score | 3,842 (90.7) | | 4,376 (90.2) | |
| ≥ 13 tot score | 392 (9.3) | | 474 (9.8) | |
| **Self-harm 17 years** | 4,183 (98.8) | 52 (1.2) | 4,758 (98.1) | 94 (1.9) |
| No | 3,470 (82.9) | | 3,932 (82.6) | |
| Yes | 713 (17.1) | | 826 (17.4) | |
| **Suicide attempt 17 years** | 4,185 (98.8) | 50 (1.2) | 4,760 (98.1) | 92 (1.9) |
| No | 4,014 (95.9) | | 4,551 (95.6) | |
| Yes | 171 (4.1) | | 209 (4.4) | |
| | **Mean (SD)** | | **Mean (SD)** | |
| **Psychological distress 17 years** | 6.13 (4.34) | 1 (0.02) | 6.13 (4.41) | 2 (0.04) |

The following rules apply to all the variables in the table:
* % uses whole sample as denominator
** % uses sample with complete data on a specific variable as denominator
N = number of participants; % = percentage of the number of participants; SD = standard deviation; PSM = propensity score matching; MI = multiple imputation.

### Table S4. Descriptive analyses – Matching & confounding variables by type of sample

| | Full sample (all the variables: both matching and confounding variables) | | | | PSM sample (only suitable matching variables) | | | |
|---|---|---|---|---|---|---|---|---|
| | **Girls** | | **Boys** | | **Girls** | | **Boys** | |
| **Matching variables** | | | | | | | | |
| | **Sample N (%)** | **Missingness N (%)** | **Sample N (%)** | **Missingness N (%)** | **Sample N (%)** | **Missingness N (%)** | **Sample N (%)** | **Missingness N (%)** |
| **Total obs.** | **5,119 (100.0)** | | **4,852 (100.0)** | | **4,547 (100.0)** | | **4,235 (100.0)** | |
| **Ethnicity** | | 0 (0.0) | | 1 (0.02) | | 0 (0) | | 0 (0) |
| White | 4,138 (80.8) | | 3,925 (80.9) | | 3,704 (81.4) | | 3,455 (81.6) | |
| Mixed | 159 (3.1) | | 136 (2.8) | | 128 (2.8) | | 120 (2.8) | |
| Indian | 143 (2.8) | | 152 (3.1) | | 129 (2.8) | | 128 (3.0) | |
| Pakistani and Bangladeshi | 418 (8.2) | | 376 (7.8) | | 369 (8.1) | | 320 (7.6) | |
| Black or Black British | 181 (3.5) | | 178 (3.7) | | 152 (3.3) | | 138 (3.2) | |
| Other ethnic group | 80 (1.6) | | 84 (1.7) | | 69 (1.5) | | 74 (1.8) | |
| **Parental education** | | 390 (7.6) | | 394 (8.1) | | 0 (0) | | 0 (0) |
| No higher education | 2,073 (43.8) | | 1,903 (42.7) | | 1,970 (43.3) | | 1,762 (41.6) | |
| Higher education | 2,656 (56.2) | | 2,555 (57.3) | | 2,581 (56.7) | | 2,473 (58.4) | |
| **Pubertal status at 14** | | 501 (9.8) | | 568 (11.7) | | 0 (0) | | 0 (0) |
| Not yet started to grow | 23 (0.5) | | 41 (1.0) | | 22 (0.5) | | 39 (0.9) | |
| Started to grow | 4,595 (99.5) | | 4,243 (99.0) | | 4,529 (99.5) | | 4,196 (99.1) | |
| **Sexual identity at 17** | | 24 (0.5) | | 14 (0.3) | | 0 (0) | | 0 (0) |
| Heterosexual | 4,357 (85.5) | | 4,527 (93.6) | | 3,889 (85.5) | | 3,973 (93.8) | |
| Other than heterosexual | 738 (14.5) | | 311 (6.4) | | 662 (14.5) | | 262 (6.2) | |
| **Self-harm at 14** | | 478 (9.3) | | 505 (10.4) | | 0 (0) | | 0 (0) |
| No | 3,648 (78.6) | | 3,991 (91.8) | | 3,571 (78.5) | | 3,900 (92.1) | |
| Yes | 993 (21.4) | | 356 (8.2) | | 980 (21.5) | | 335 (7.9) | |
| **Psychological distress at 14** | | 482 (9.4) | | 528 (10.9) | | 0 (0) | | 0 (0) |
| < 12 score | 3,599 (77.6) | | 3,972 (91.9) | | 3,535 (77.7) | | 3,896 (92.0) | |
| ≥ 12 score | 1,038 (22.4) | | 352 (8.1) | | 1,016 (22.3) | | 339 (8.0) | |
| **Sexual violence reported at age 14** | | 458 (8.9) | | 508 (10.5) | | | | |
| No | 4,462 (95.7) | | 4,289 (98.7) | | 4,157 (98.8) | 0 (0) | 4,157 (98.8) | 0 (0) |
| Yes | 199 (4.3) | | 55 (1.3) | | 51 (1.2) | | 51 (1.2) | |

| | Mean (SD) | | Mean (SD) | | Mean (SD) | | Mean (SD) | |
|---|---|---|---|---|---|---|---|---|
| **Psychological distress at 14** | 6.99 (6.53) | 482 (9.4) | 4.05 (4.57) | 528 (10.9) | 6.97 (6.53) | 0 (0) | 4.02 (4.5) | 0 (0) |
| **Confounding variables** | | | | | | | | |
| **Family income** | | 381 (7.4) | | 386 (8.0) | | | | |
| 1 | 737 (15.6) | | 680 (15.2) | | | | | |
| 2 | 761 (16.1) | | 664 (14.9) | | | | | |
| 3 | 944 (19.9) | | 896 (20.1) | | | | | |
| 4 | 1,117 (23.6) | | 1,109 (24.8) | | | | | |
| 5 | 1,179 (24.9) | | 1,117 (25.0) | | | | | |
| **Single parent household** | | 376 (7.4) | | 380 (7.8) | | | | |
| Two parents/carers | 3,690 (77.8) | | 3,495 (78.2) | | | | | |
| One parent/carer | 1,053 (22.2) | | 977 (21.8) | | | | | |
| **Age of menarche** | | 1,062 (20.8) | | | | | | |
| < 11 years | 223 (5.5) | | N/A | | | | | |
| ≥ 11 years | 3,834 (94.5) | | N/A | | | | | |
| **Early sexual activity** | | 488 (9.5) | | 556 (11.5) | | | | |
| No | 4,270 (92.2) | | 3,850 (89.6) | | | | | |
| Yes | 361 (7.8) | | 446 (10.4) | | | | | |
| **Relationship status** | | 456 (8.9) | | 512 (10.5) | | | | |
| No | 3,931 (84.3) | | 3,599 (82.9) | | | | | |
| Yes | 732 (15.7) | | 741 (17.1) | | | | | |
| **Peer relationship quality** | | 430 (8.4) | | 435 (9.0) | | | | |
| Has no close friends | 100 (2.1) | | 170 (3.8) | | | | | |
| Has close friends | 4,589 (97.9) | | 4,247 (96.2) | | | | | |
| **Having been bullied** | | 450 (8.8) | | 497 (10.2) | | | | |
| Never | 878 (18.8) | | 1,180 (27.1) | | | | | |
| Sometimes | 2,178 (46.7) | | 1,991 (45.7) | | | | | |
| Often | 1,613 (34.5) | | 1,184 (27.2) | | | | | |
| **Missed school** | | 435 (8.5) | | 441 (9.1) | | | | |
| Never | 4,301 (91.8) | | 4,052 (91.9) | | | | | |
| Sometimes | 303 (6.5) | | 307 (7.0) | | | | | |
| Yes | 80 (1.7) | | 52 (1.2) | | | | | |

| | | | | |
|---|---|---|---|---|
| **Risky behaviours** | | | | |
| *Ever smoked* | | 473 (9.2) | | 517 (10.7) |
| No | 3,986 (85.8) | | 3,832 (88.4) | |
| Yes | 660 (14.2) | | 503 (11.6) | |
| *Ever consumed alcohol* | | 459 (9.0) | | 502 (10.4) |
| No | 2,716 (58.3) | | 2,437 (56.0) | |
| Yes | 1,944 (41.7) | | 1,913 (44.0) | |
| *Heavy alcohol freq.* | | 464 (9.1) | | 515 (10.6) |
| Never | 2,716 (58.4) | | 2,437 (56.2) | |
| Sometimes | 1,570 (33.7) | | 1,568 (36.1) | |
| Often | 228 (4.9) | | 237 (5.5) | |
| Very often | 141 (3.0) | | 95 (2.2) | |
| *Ever taken drugs* | | 454 (8.9) | | 502 (10.4) |
| No | 4,490 (96.2) | | 4,178 (96.0) | |
| Yes | 175 (3.8) | | 172 (4.0) | |
| **Lifelong disability/illness** | | 423 (8.3) | | 411 (8.5) |
| No | 3,916 (83.4) | | 3,580 (80.6) | |
| Yes | 780 (16.6) | | 861 (19.4) | |
| | **Mean (SD)** | | **Mean (SD)** | |
| **Number of siblings** | 1.55 (1.15) | 376 (7.4) | 1.56 (1.14) | 379 (7.8) |
| **BMI** | 21.92 (4.16) | 633 (12.4) | 20.87 (3.97) | 495 (10.2) |
| **Life satisfaction** | 26.02 (6.84) | 452 (8.8) | 28.17 (6.14) | 499 (10.3) |

N = number of participants; % = percentage of the number of participants; SD = standard deviation; PSM = propensity score matching; MI = multiple imputation.

### Table S5. PSM – Key matching variables (before and after matching)

| | Girls | | | Boys | | |
|---|---|---|---|---|---|---|
| | **Sexual violence group** | **Unmatched control group** | **Matched control group (all variables)** | **Sexual violence group** | **Unmatched control group** | **Matched control group (all variables)** |
| | **N (%)** | **N (%)** | **N (%)** | **N (%)** | **N (%)** | **N (%)** |
| **Group sample** | 938 (20.6) | 3,609 (79.4) | 938 (50.0) | 230 (5.4) | 4,005 (94.6) | 230 (50.0) |
| **Ethnicity** | | | | | | |
| White | 797 (85.0) | 2,904 (80.5) | 828 | 202 (87.8) | 3,253 (81.2) | 206 |
| Mixed | 34 (3.6) | 94 (2,6) | 34 | 8 (3.5) | 112 (2.8) | 8 |
| Indian | 25 (2.7) | 104 (2.9) | 19 | 4 (1.7) | 124 (3.1) | 4 |
| Pakistani and Bangladeshi | 41 (4.4) | 328 (9.1) | 31 | 5 (2.2) | 315 (7.9) | 5 |
| Black or Black British | 26 (2.8) | 125 (3.5) | 20 | 7 (3.0) | 131 (3.3) | 5 |
| Other | 15 (1.6) | 54 (1.5) | 6 | 4 (1.7) | 70 (1.8) | 2 |
| **Parental education** | | | | | | |
| No higher education | 341 (36.4) | 1,628 (45.1) | 335 | 74 (32.2) | 1,688 (42.2) | 77 |
| Higher education | 597 (63.6) | 1,981 (54.9) | 603 | 156 (67.8) | 2,317 (57.9) | 153 |
| **Pubertal status at 14** | | | | | | |
| Not yet started to grow | 1 (0.1) | 21 (0.6) | 0 | 1 (0.4) | 38 (1.0) | 1 |
| Started to grow | 937 (99.9) | 3,588 (99.4) | 938 | 229 (99.6) | 3,967 (99.0) | 229 |
| **Sexual identity at 17** | | | | | | |
| Heterosexual | 724 (77.2) | 3,163 (87.6) | 723 | 183 (80.0) | 3,790 (94.6) | 187 |
| Other than heterosexual | 214 (22.8) | 446 (12.4) | 215 | 47 (20.4) | 215 (5.4) | 43 |
| **Self-harm at 14** | | | | | | |
| No | 598 (63.7) | 2,969 (82.3) | 597 | 187 (81.3) | 3,713 (92.7) | 186 |

| | | | | | | |
|---|---|---|---|---|---|---|
| Yes | 340 (36.3) | 640 (17.7) | 341 | 43 (18.7) | 292 (7.3) | 44 |
| **Psychological distress at 14** | | | | | | |
| < 12 score | 600 (64.0) | 2,932 (81.2) | 581 | 192 (83.5) | 3,704 (92.5) | 186 |
| ≥ 12 score | 338 (36.0) | 677 (18.8) | 357 | 38 (16.5) | 301 (7.5) | 44 |
| **Sexual violence <14** | | | | | | |
| No | 834 (88.9) | 3,514 (97.4) | 858 | 220 (95.6) | 3,964 (99.0) | 219 |
| Yes | 104 (11.1) | 95 (2.6) | 80 | 10 (4.4) | 41 (1.0) | 11 |
| | **Mean (SE)** | **Mean (SE)** | **Mean (SE)** | **Mean (SE)** | **Mean (SE)** | **Mean (SE)** |
| **Psychological distress at 14** | 9.57 (0.24) | 6.30 (0.10) | 9.82 (0.24) | 6.06 (0.38) | 3.90 (0.07) | 6.42 (0.42) |

PSM = propensity score matching.

### Table S6a. Matching quality evaluation for all the matching variables (girls) – PSM

| | Mean | | | t-test | |
|---|---|---|---|---|---|
| **Matching variables** | **Treated** | **Control** | **% bias** | **t** | **p>\|t\|** |
| Ethnicity | | | | | |
| Mixed | 0.03625 | 0.03625 | 0 | 0 | 1 |
| Indian | 0.02665 | 0.02026 | 3.9 | 0.92 | 0.36 |
| Pakistani and Bangladeshi | 0.04371 | 0.03305 | 4.3 | 1.2 | 0.23 |
| Black or Black British | 0.02772 | 0.02132 | 3.7 | 0.9 | 0.371 |
| Other | 0.01599 | 0.0064 | 7.8 | 1.98 | 0.048 |
| Parental education | 0.63646 | 0.64286 | -1.3 | -0.29 | 0.773 |
| Pubertal status | 0.99893 | 1 | -1.8 | -1 | 0.317 |
| Sexual identity | 0.22814 | 0.22921 | -0.3 | -0.05 | 0.956 |
| Self-harm at 14 | 0.36247 | 0.36354 | -0.2 | -0.05 | 0.962 |
| Depressive symptoms at 14 (binary) | 0.36034 | 0.3806 | -4.6 | -0.91 | 0.364 |
| Depressive symptoms at 14 (continuous) | 9.5693 | 9.8177 | -3.7 | -0.74 | 0.458 |
| Sexual violence <14 | 0.11087 | 0.08529 | 10.3 | 1.86 | 0.063 |

PSM = propensity score matching. % percentage.

### Table S6b. Matching quality evaluation for all the matching variables (boys) – PSM

| | Mean | | | t-test | |
|---|---|---|---|---|---|
| **Matching variables** | **Treated** | **Control** | **% bias** | **t** | **p>\|t\|** |
| Ethnicity | | | | | |
| Mixed | 0.03478 | 0.03478 | 0 | 0 | 1 |
| Indian | 0.01739 | 0.01739 | 0 | 0 | 1 |
| Pakistani and Bangladeshi | 0.02174 | 0.02174 | 0 | 0 | 1 |
| Black or Black British | 0.03043 | 0.02174 | 5 | 0.58 | 0.56 |
| Other | 0.01739 | 0.0087 | 6.6 | 0.82 | 0.412 |
| Parental education | 0.67826 | 0.66522 | 2.7 | 0.3 | 0.766 |
| Pubertal status | 0.99565 | 0.99565 | 0 | 0 | 1 |
| Sexual identity | 0.20435 | 0.18696 | 5.3 | 0.47 | 0.639 |
| Self-harm at 14 | 0.18696 | 0.1913 | -1.3 | -0.12 | 0.905 |
| Depressive symptoms at 14 (binary) | 0.16522 | 0.1913 | -8.1 | -0.73 | 0.466 |
| Depressive symptoms at 14 (continuous) | 6.0565 | 6.4174 | -7 | -0.64 | 0.525 |
| Sexual violence <14 | 0.04348 | 0.04783 | -2.7 | -0.22 | 0.824 |

PSM = propensity score matching. % percentage.

**Figure S3a. Graph of the matching quality evaluation for all the matching variables (girls)**

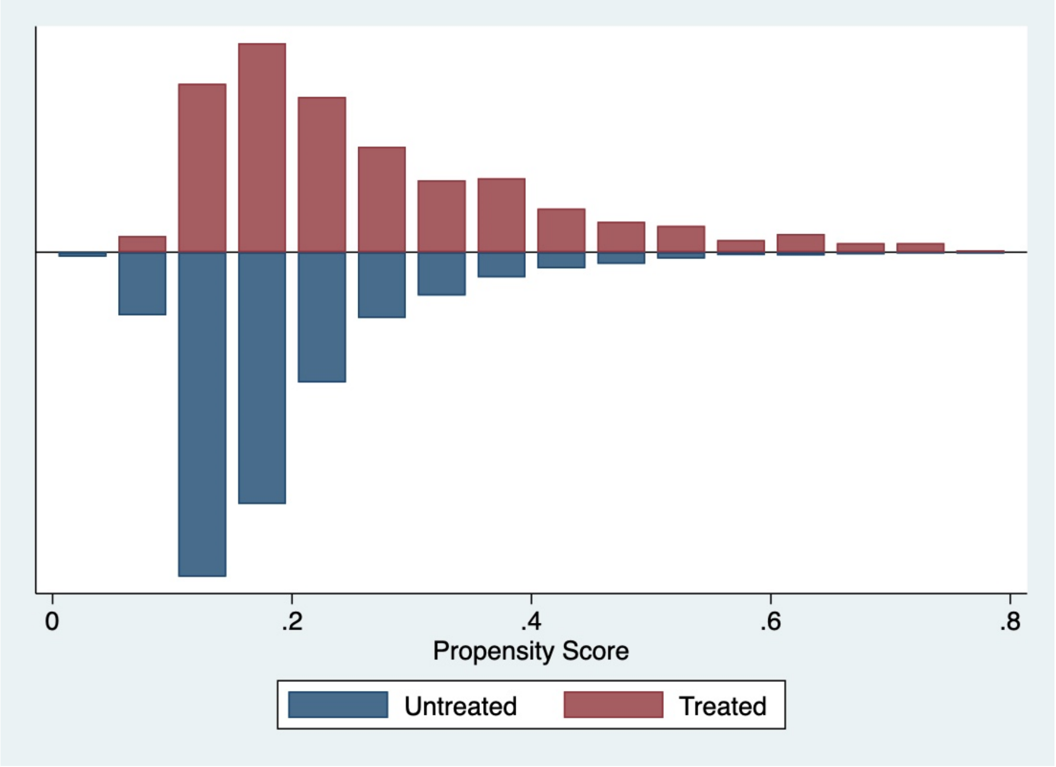

**Figure S3b. Graph of the matching quality evaluation for all the matching variables (boys)**

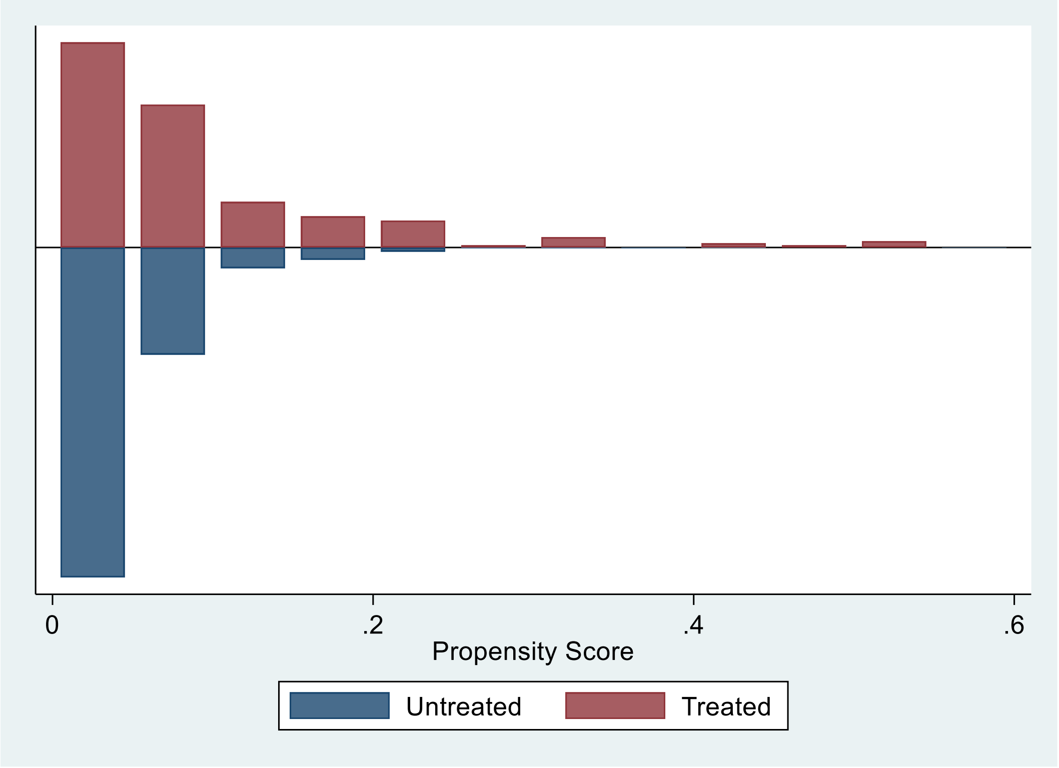

**Table S7. Regression coefficients and risk ratios for sexual violence – PSM analysis excluding those who reported sexual violence before age 14 years**

| | Psychological distress (continuous) Coefficient [95% CI] | | High psychological distress RR [95% CI] | | Self-harm RR [95% CI] | | Attempted Suicide RR [95% CI] | |
|---|---|---|---|---|---|---|---|---|
| | **girls** | **boys** | **girls** | **boys** | **girls** | **boys** | **girls** | **boys** |
| **Matched samples** | | | | | | | | |
| PSM (all matching variables) | 1·52 [1·02, 2·02] | 2·13 [1·21, 3·05] | 1·39 [1·20, 1·61] | 1·63 [1·12, 2·38] | 1·55 [1·37, 1·74] | 2·19 [1·63, 2·95] | 1·54 [1·21, 1·96] | 1·67 [0·99, 2·82] |
| PSM (all minus sexual identity, pubertal status, parental education) | 2·03 [1·54, 2·52] | 2·83 [1·95, 3·71] | 1·58 [1·35, 1·84] | 2·48 [1·58, 3·88] | 1·72 [1·52, 1·95] | 2·73 [1·96, 3·78] | 1·47 [1·16, 1·86] | 2·78 [1·47, 5·23] |
| PSM (all minus psychological distress and self-harm at age 14) | 2·54 [2·06, 3·03] | 2·61 [1·72, 3·50] | 1·86 [1·58, 2·20] | 1·97 [1·31, 2·95] | 1·79 [1·57, 2·03] | 2·59 [1·88, 3·56] | 1·88 [1·45, 2·44] | 2·57 [1·39, 4·76] |
| **Matched sample with multivariable confounder adjustment** | | | | | | | | |
| Adjusted regression analysis | 1·43 [0·97, 1·89] | 1·69 [0·79, 2·60] | 1·38 [1·20, 1·59] | 1·46 [0·99, 2·15] | 1·51 [1·34, 1·70] | 2·12 [1·57, 2·87] | 1·51 [1·20, 1·91] | 1·56 [0·89, 2·74] |

RR risk ratio; CI confidence interval. PSM= Propensity score matched. * Those who reported unwelcome sexual approach and no sexual assault

Adjustment 1: prior mental health (depressive symptoms and self-harm until 14 years). Adjustment 2: sociodemographic/economic characteristics (ethnicity, parental education, family income, sexual identity, single parent household, number of siblings). Adjustment 3: puberty-related characteristics (pubertal status, age of menarche (girls), early sexual activity, sexual violence until 14 years). Adjustment 4: interpersonal characteristics (relationship status, peer relationships, bullying). Fully adjusted model: health-related characteristics (BMI, risky behaviours, missing school without parents' permission, disability, life satisfaction). As can be seen in the table there is a substantial impact on the estimates of some confounder adjustment with later adjustment having less impact on the estimates.

### Table S8. Population attributable fractions for sexual violence and mental health outcomes

| | High psychological distress<br>Estimate [CIs] | | Self-harm<br>Estimate [CIs] | | Attempted Suicide<br>Estimate [CIs] | |
|---|---|---|---|---|---|---|
| | **Girls** | **Boys** | **Girls** | **Boys** | **Girls** | **Boys** |
| **Overall sexual violence exposure** | | | | | | |
| PAF | 14.0% [9.4, 18.3] | 3.7% [-0.6, 7.9] | 16.8% [12.2, 21.1] | 8.4% [4.9, 11.8] | 18.7% [8.4, 27.9] | 10.5% [3.4, 1.7] |
| Scenario as observed | 22.6% [21.0, 24.4] | 10.2% [8.9, 11.7] | 28.9% [26.8, 31.2] | 20.3% [18.3, 22.5] | 11.0% [10.0, 12.7] | 4.3% [3.6, 5.2] |
| Scenario zero sexual violence | 19.5% [17.7, 21.5] | 9.8% [8.5, 11.4] | 24.0% [21.7, 26.7] | 18.6% [16.5, 20.9] | 9.1% [7.8, 10.8] | 3.9% [3.2, 4.7] |
| **Sexual assault exposure** | | | | | | |
| PAF | 4.1% [1.9, 6.2] | 2.0% [-0.4, 4.4] | 5.6% [3.8, 7.4] | 1.9% [0.9, 2.9] | 8.1% [4.3, 11.8] | 5.5% [0.6, 10.1] |
| Scenario as observed | 22.6% [21.0, 24.4] | 10.2% [8.9, 11.7] | 28.9% [26.8, 31.2] | 20.3% [18.2, 22.6] | 11.0% [9.9, 12.7] | 4.3% [3.6, 5.2] |
| Scenario zero sexual assault | 21.7% [20.1, 23.5] | 10.0% [8.7, 11.5] | 27.0% [25.1, 29.6] | 19.9% [17.8, 22.3] | 10.3% [9.0, 11.8] | 4.1% [3.4, 4.9] |
| **Unwelcome sexual approach exposure** | | | | | | |
| PAF | 12.6% [8.1, 16.8] | 3.7% [-0.6, 7.9] | 15.7% [11.3, 19.8] | 8.2% [4.7, 11.6] | 16.5% [6.5, 25.5] | 1.0% [3.3, 17.0] |
| Scenario as observed | 22.6% [21.0, 24.4] | 10.2% [8.9, 11.7] | 28.9% [26.8, 31.2] | 20.3% [18.3, 22.5] | 11.3% [10.0, 12.7] | 4.3% [3.6, 5.2] |
| Scenario zero unwanted sexual approach | 19.8% [18.0, 21.8] | 9.8% [8.5, 11.4] | 24.4% [22.1, 27.0] | 18.6% [16.6, 20.9] | 9.4% [8.0, 11.0] | 3.9% [3.2, 4.7] |

PAF population attributable fraction; CI confidence interval. Note: these models are based on the full analytic sample and fully adjusted multivariable models.

### MeSH Terms used in literature search in PubMed

**Sexual violence**

- Offense, Sex
- Offences, Sex
- Sex Offense
- Sexual Assault
- Assault, Sexual
- Assaults, Sexual
- Sexual Assaults
- Sexual Violence
- Sexual Violences
- Violence, Sexual
- Violences, Sexual
- Sexual Abuse
- Abuse, Sexual
- Abuses, Sexual
- Sexual Abuses
- Sexual Harassment
- Harassment, Sexual
- Harassments, Sexual
- Sexual Harassments
- Sexual Harrassment
- Harrassment, Sexual
- Harrassments, Sexual
- Sexual Harrassments

**Mental health**

- Psychological Distress
- Distress, Psychological
- Emotional Distress
- Distress, Emotional
- Emotional Stress
- Stress, Emotional
- Depression
- Depressive Symptoms
- Depressive Symptom
- Symptom, Depressive
- Symptoms, Depressive
- Emotional Depression
- Depression, Emotional
- Behavior, Self-Injurious
- Self Injurious Behavior
- Self-Injurious Behaviors
- Intentional Self Injury
- Intentional Self Injuries
- Self Injury, Intentional
- Intentional Self Harm
- Self Harm, Intentional
- Nonsuicidal Self Injury
- Nonsuicidal Self Injuries
- Self Injury, Nonsuicidal
- Deliberate Self-Harm
- Deliberate Self Harm

- Self-Harm, Deliberate
- Self-Injury
- Self Injury
- Non-Suicidal Self Injury
- Non Suicidal Self Injury
- Non-Suicidal Self Injuries
- Self Injury, Non-Suicidal
- Self Harm
- Harm, Self
- Self-Destructive Behavior
- Behavior, Self-Destructive
- Self Destructive Behavior
- Self-Destructive Behaviors
- Attempted Suicide
- Suicide Attempt
- Attempt, Suicide
- Parasuicide
- Parasuicides

**Population**

- Adolescents
- Adolescence
- Teens
- Teen
- Teenagers
- Teenager
- Youth
- Youths
- Adolescents, Female
- Adolescent, Female
- Female Adolescent
- Female Adolescents
- Adolescents, Male
- Adolescent, Male
- Male Adolescent
- Male Adolescents

### References

1. Butler AC. Child sexual assault: Risk factors for girls. Child Abuse Negl. 2013;37(9):643–52.

2. Landstedt E, Gillander Gådin K. Deliberate self-harm and associated factors in 17-year-old Swedish students. Scandinavian Journal of Public Health. 2011;39(1):17–25.

3. Coulter RWS, Mair C, Miller E, Blosnich JR, Matthews DD, McCauley HL. Prevalence of Past-Year Sexual Assault Victimization Among Undergraduate Students: Exploring Differences by and Intersections of Gender Identity, Sexual Identity, and Race/Ethnicity. Prev Sci [Internet]. 2017 Aug;18(6):726–36. Available from: https://pubmed.ncbi.nlm.nih.gov/28210919

4. Moore L, Jayaweera H, Redshaw M, Quigley M. Migration, ethnicity and mental health: evidence from mothers participating in the Millennium Cohort Study. Public Health. 2019;171:66–75.

5. Plöderl M, Tremblay P. Mental health of sexual minorities. A systematic review. International review of psychiatry. 2015;27(5):367–85.

6. Barrett AE, Turner RJ. Family structure and mental health: The mediating effects of socioeconomic status, family process, and social stress. J Health Soc Behav. 2005;46(2):156–69.

7. Finkelhor D, Ormrod RK, Turner HA. Re-victimization patterns in a national longitudinal sample of children and youth. Child Abuse and Neglect. 2007;31(5):479–502.

8. Lawson DW, Mace R. Siblings and childhood mental health: Evidence for a later-born advantage. Social Science & Medicine. 2010;

9. Mendle J, Ryan RM, Mckone KM. Early Childhood Maltreatment and Pubertal Development: Replication in a Population-Based Sample. 2015;

10. Vicary JR, Klingaman LR, Harkness WL. Risk factors associated with date rape and sexual assault of adolescent girls. J Adolesc. 1995;18(3):289–306.

11. Organization WH. Global and regional estimates of violence against women: prevalence and health effects of intimate partner violence and non-partner sexual violence. World Health Organization; 2013.

12. Coggan C, Bennett S, Hooper R, Dickinson P. International Journal of Mental Health Promotion Association between Bullying and Mental Health Status in New Zealand Adolescents E E Association between Bullying and Mental Health Status in New Zealand Adolescents. International Journal of Mental Health Promotion VOLUME. 2003;5(1):16–22.

13. DUNCAN RD. Peer and Sibling Aggression:: An Investigation of Intra- and Extra-Familial Bullying. Journal of Interpersonal Violence. 1999 Aug 1;14(8):871–86.

14. Aucott L. Mental Well-Being Related To Lifestyle and Risky Behaviours in 18-25 Year Old: Evidence from North-East Scotland. International Journal of Public Health Research. 2014 Mar 1;4(1 SE-Public Health Research Articles).

15. Charak R, Koot HM, Dvorak RD, Elklit A, Elhai JD. Unique versus cumulative effects of physical and sexual assault on patterns of adolescent substance use. Psychiatry Research. 2015 Mar 5;230(3):763–9.

16. RUNTZ M, BRIERE J. Adolescent “Acting-Out” and Childhood History of Sexual Abuse. Journal of Interpersonal Violence. 1986 Sep 1;1(3):326–34.

17. Choudhary E, Coben JH, Bossarte RM. Gender and time differences in the associations between sexual violence victimization, health outcomes, and risk behaviors. Am J Mens Health. 2008;2(3):254–9.

18. Alriksson-Schmidt AI, Armour BS, Thibadeau JK. Are adolescent girls with a physical disability at increased risk for sexual violence? Journal of School Health. 2010;80(7):361–7.

19. Kessler RC, Andrews G, Colpe LJ, Hiripi E, Mroczek DK, Normand SL, et al. Short screening scales to monitor population prevalences and trends in non-specific psychological distress. Psychol Med. 2002;32(6):959–76.

20. Vissoci JRN, Vaca SD, El-Gabri D, de Oliveira LP, Mvungi M, Mmbaga BT, et al. Cross-cultural adaptation and psychometric properties of the Kessler Scale of Psychological Distress to a traumatic brain injury population in Swahili and the Tanzanian Setting. Health and Quality of Life Outcomes. 2018;16(1):1–8.

21. Jang Y, Powers DA, Yoon H, Rhee MK, Park NS, Chiriboga DA. Measurement equivalence of English versus native language versions of the Kessler 6 (K6) Scale: An examination in three Asian American groups. Asian Am J Psychol. 2018;9(3):211.

22. Ma M, Fang J, Li C, Bao J, Zhang Y, Chen N, et al. The status and high risk factors of severe psychological distress in migraine patients during nCOV-2019 outbreak in Southwest China: a cross-sectional study. The Journal of Headache and Pain. 2020;21(1):1–7.

23. Furukawa TA, Kawakami N, Saitoh M, Ono Y, Nakane Y, Nakamura Y, et al. The performance of the Japanese version of the K6 and K10 in the World Mental Health Survey Japan. Int J Methods Psychiatr Res. 2008;17(3):152–8.

24. Easton SD, Safadi NS, Wang Y, Hasson RG. The Kessler psychological distress scale: translation and validation of an Arabic version. Health Qual Life Outcomes. 2017;15(1):1–7.

25. Kawakami N, Thi Thu Tran T, Watanabe K, Imamura K, Thanh Nguyen H, Sasaki N, et al. Internal consistency reliability, construct validity, and item response characteristics of the Kessler 6 scale among hospital nurses in Vietnam. PLoS One. 2020;15(5):e0233119.

26. Staples LG, Dear BF, Gandy M, Fogliati V, Fogliati R, Karin E, et al. Psychometric properties and clinical utility of brief measures of depression, anxiety, and general distress: The PHQ-2, GAD-2, and K-6. Gen Hosp Psychiatry. 2019;56:13–8.

27. Messer SC, Angold A, Costello EJ, Loeber R, al et. Development of a short questionnaire for use in epidemiological studies of depression in children and adolescents: Factor composition and structure across development. International Journal of Methods in Psychiatric Research. 1995;5(4):251–62.

28. Lundervold AJ, Breivik K, Posserud MB, Stormark KM, Hysing M. Symptoms of depression as reported by Norwegian adolescents on the Short Mood and Feelings Questionnaire. Front Psychol. 2013;4:613.

29. Mori I. Millennium Cohort Study Seventh Sweep (MCS7) Technical Report. 2019.
